# Altered Semantic Prediction Error Processing with Increasing Schizotypal and Autistic Traits

**DOI:** 10.64898/2026.01.09.26343667

**Authors:** Elisabeth F. Sterner, Verena F. Demler, Rachel Nuttall, Lucy J. MacGregor, Christoph Mathys, Franziska Knolle

## Abstract

Predictive processing has been proposed as an explanatory framework for symptom development in both autism (ASD) and schizophrenia (SSD) spectrum disorders, with ASD being associated with an overweighting of (low-level) sensory evidence whereas SSD is characterized by an overweighting of (high-level) prior beliefs. The goal of the present study was to investigate these hypotheses in subclinical expressions of ASD and SSD in the domain of language processing. To test this, we used an auditory comprehension task designed to directly manipulate the precision of high-level semantic prior beliefs and low-level sensory evidence. We applied hierarchical Bayesian belief updating modeling to quantify this effect and used EEG to examine whether an imbalance in the weighting of prior beliefs and sensory evidence would be characterized by altered processing of semantic precision-weighted prediction errors as indexed by alterations in mean N400 amplitudes. Computational modeling revealed that increasing schizotypal traits were associated with a significant overweighting of prior beliefs, while autistic traits did not show a significant shift. Linear mixed models on the mean N400 amplitudes further indicated that this schizotypy-related overweighting of semantic prior beliefs was reflected in a reduced semantic prediction error signal, indexed by smaller N400 differences between low entropy sentences and both high and low-mismatch sentences. A similar pattern emerged for increasing autistic traits, though the effect was weaker and less distinct, pointing to a subtle overweighting of semantic prior beliefs, only. Overall, our findings provide converging computational and electrophysiological support for an overweighting of semantic prior beliefs with increasing subclinical schizotypy, consistent with predictive processing accounts of SSD, whereas we did not find evidence for an overweighting of sensory evidence with increasing autistic traits, with electrophysiological results instead pointing toward subtle alterations in the weighting of semantic prior beliefs.

## INTRODUCTION

Schizophrenia (SSD) and autism spectrum disorder (ASD) are distinct psychiatric diagnoses that, despite differences in onsets, clinical presentation, and long-term trajectories, intersect regarding their genetic predisposition, environmental risk factors, and symptom features(Jutla et al., 2022; Zheng et al., 2018). The overlap is further supported epidemiologically with increased prevalence of cooccurring diagnoses of schizophrenia in autism and vice versa(Lai et al., 2019; Zheng et al., 2018) and is also reflected in a strong correlation of subclinical schizotypal and autistic personality traits in the general population(Nenadić et al., 2021; Zhou et al., 2019). For explaining this close relationship between SSD and ASD, predictive processing has been put forward as a promising framework among various theoretical accounts(Chisholm et al., 2015; Nenadić et al., 2021), providing insights into the computational similarities and differences between the conditions. Predictive processing suggests that the brain performs Bayesian inference to respond efficiently to its environment by integrating high-level prior beliefs generated from an internal model of the world with incoming low-level sensory information(Friston, 2005). During this inferential process, prior beliefs and sensory evidence are weighted according to their respective precision (i.e. inverse variance) with those signals with higher precision having a greater impact on the posterior belief, which corresponds to the perception. The discrepancy between prior beliefs and sensory evidence, defined as the precision-weighted prediction error, drives updates of the internal model (**Figure 1A & 1B**) to generate more precise prior beliefs in the future. In this framework, symptom development in both SSD(Adams et al., 2013; Fletcher & Frith, 2009; Goodwin et al., 2025; Harding et al., 2024; Sterzer et al., 2018, 2019) and ASD(Haker et al., 2016; Lawson et al., 2014; Pellicano & Burr, 2012; van Schalkwyk et al., 2017) has been linked to disruptions in the weighting process of prior beliefs relative to sensory evidence. However, the exact mechanisms underlying this imbalance remain under debate.

**Figure 1.**
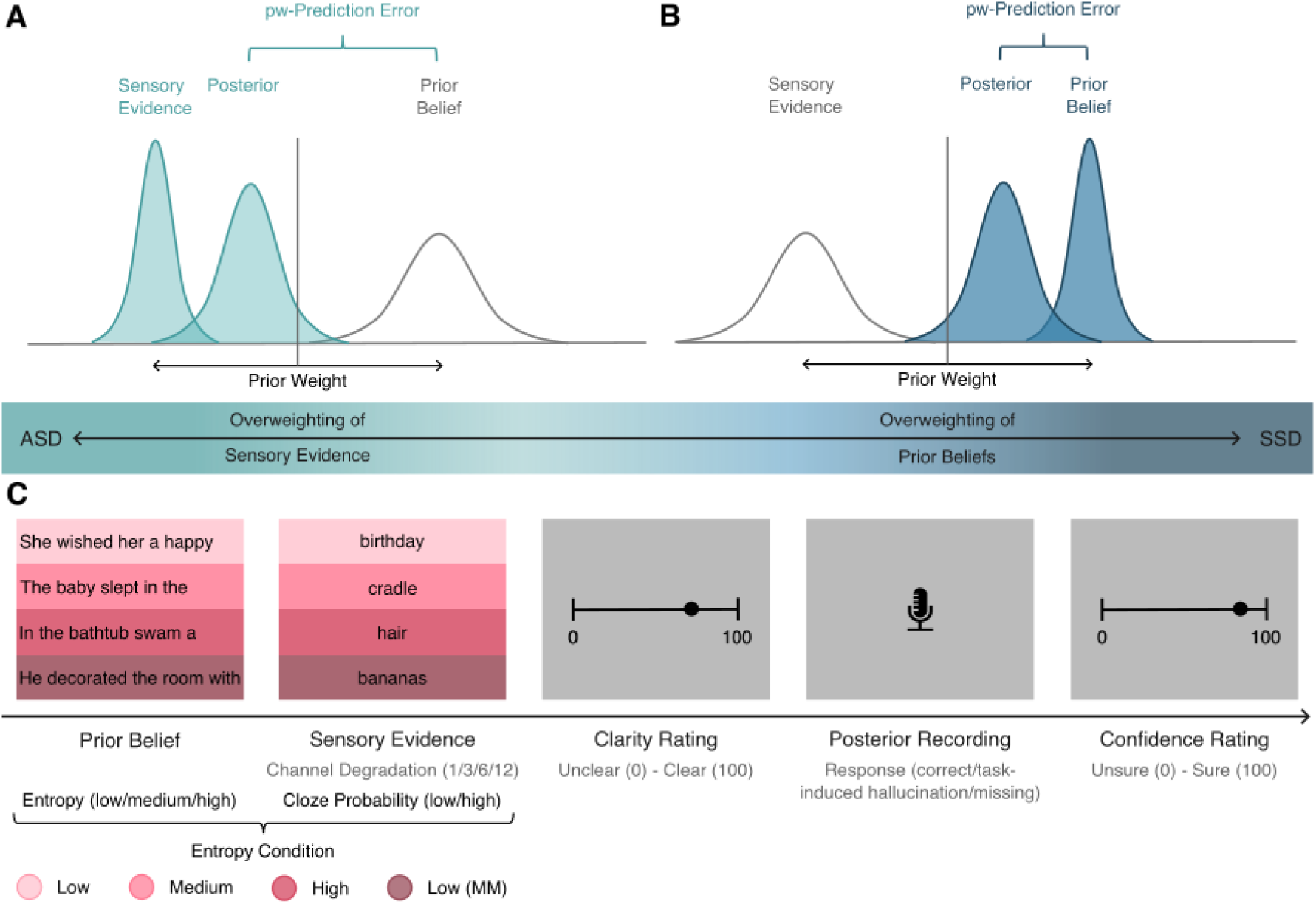
**A) & B) ASD-SSD Predictive Coding Spectrum**. A) shows an overweighting of sensory evidence and resulting in increased prediction error signaling (visualized as the difference between the prior beliefs and the posterior) as suggested in ASD, whereas B) displays an overweighting of prior beliefs with attenuated prediction error signaling as suggested in SSD. In both A) and B) the posterior moves towards the more strongly weighted component indicated by higher precision, i.e., lower variance of the distribution. **C) Predictive Language Processing Task.** On each trial, participants listened to sentences via headphones. High-level semantic prior beliefs with varying precisions were induced by sentence beginnings with three predictability levels measured in entropy (low, N=80; medium, N=80; high, N=40; from light pink to pink). Half of the low entropy sentences (N=40) were completed by a sentence-final word with a cloze probability of 0, forming the Low (Mismatch; MM) condition (dark pink). The remaining 160 sentences were completed with the highest cloze probability response as assessed in an independent sample. The precision of the sensory evidence was further modulated by the auditory clarity of the sentence-final words, which were presented in four different clarity levels (using noise vocoding with 1, 3, 6, 12 channels ranging from unintelligible to highly intelligible). After the presentation of the final word, participants rated the clarity of the target word (Likert scale: 0-100). Participants were then asked to say the word they heard and rate how confident they were with their response (Likert scale: 0-100).

In ASD, low-level sensory information appears to be overweighted(Karvelis et al., 2018; Tarasi et al., 2023; Zaidel et al., 2015), giving rise to prediction errors that are disproportionately driven by misassigned sensory precision and salience(Balsters et al., 2017; Kinard et al., 2020; Mosner et al., 2019; van Laarhoven et al., 2020), and that may contribute to the feeling of sensory overload (**Figure 1A**). Persistent prediction errors may propagate up the hierarchical system, leading to increased uncertainty at higher hierarchical levels, which may in turn explain difficulties in forming generalizable representations in ASD(Lawson et al., 2014; Pellicano & Burr, 2012). The reduced influence of prior beliefs(Amoruso et al., 2019; Eckert et al., 2024; Ganglmayer et al., 2020; Lieder et al., 2019; Sevgi et al., 2020) may contribute to social and communicative difficulties, such as a tendency toward a literal language comprehension(Haker et al., 2016) as well as behavioral inflexibility and specific interests. These behaviors may in turn function as compensatory strategies to minimize prediction error signaling and reduce cognitive uncertainty(van Schalkwyk et al., 2017).

Like ASD, SSD has been associated with a relative overweighting of low-level sensory information(Eckert et al., 2023; Goodwin et al., 2023; Stuke et al., 2019; Weilnhammer et al., 2020) and abnormal prediction error signaling(Cole et al., 2020; Ermakova et al., 2018; Katthagen et al., 2020), which may give rise to delusional beliefs(A. Powers et al., 2025). In contrast to ASD, SSD is additionally characterized by an increased weighting of high-level prior beliefs(Cassidy et al., 2018; Haarsma et al., 2020; Knolle et al., 2024; A. R. Powers et al., 2017; Stuke et al., 2021; Teufel et al., 2015) (**Figure 1B**). This imbalance may account for the persistence of delusions and their resistance to contradictory evidence, ultimately resulting in an inappropriate and fixed world model(Corlett et al., 2010; Sterzer et al., 2018). Moreover, overweighted prior beliefs may override incoming sensory input, giving rise to hallucinations.

These distinct predictive processing profiles of ASD and SSD have led to the proposal of an autism-schizophrenia predictive processing continuum, on with ASD is defined by a relative overweighting of (low-level) sensory information and SSD by a relative overweighting of (high-level) prior beliefs (**Figure 1A&B**)(Tarasi et al., 2022; van Schalkwyk et al., 2017). While this conceptualization might be useful for characterizing the relationship between the two conditions, only a small number of studies have tested hypotheses using experimental paradigms in shared cohorts or computational modelling(Eckert et al., 2024; Karvelis et al., 2018) or examined neurobiological correlates using imaging methods(Tarasi et al., 2023).

The present study aims to address these limitations by focusing on semantic language processing, a core impairment in both conditions(Groen et al., 2008; Kuperberg, 2010a, 2010b; Tager-Flusberg et al., 2005), which is also linked to broader phenomenology, including verbal hallucinations in SSD or communication difficulties in ASD. Beyond its clinical relevance, language processing which requires dynamic weighting of prior knowledge (i.e., contextual information) with sensory input (i.e., the speech signal)(Kuperberg & Jaeger, 2016; Ryskin & Nieuwland, 2023) to allow for accurate comprehension even under challenging conditions (i.e., noisy environments), provides a valuable framework for studying predictive processing mechanisms. When this process is disrupted – either due to an overweighting of incoming sensory information, as suggested in ASD, or an overweighting of high-level prior beliefs, as proposed in SSD – language processing may become biased, resulting in misperceptions. Building on this approach, we designed an auditory predictive language task in which we manipulated the strength of semantic prior beliefs by presenting sentences with varying levels of predictability, and varied the precision of sensory evidence, by altering auditory clarity and surprisal of sentence-final words(Knolle et al., 2024). Using a Bayesian belief updating model, we found that in two independent samples (N_1_=109, N_2_=55) higher schizotypal traits were associated with an overweighting of semantic prior beliefs and a higher frequency of misperceptions.

In this study, we aim to extend these findings by comparing potential predictive language processing impairments in individuals with varying levels of schizotypal and autistic traits, and importantly, investigate how potential computational alterations underlying these impairments manifest at a neurophysiological level.

Therefore, we combined our experimental paradigm with electroencephalography (EEG), focusing on the N400 component, an event-related potential (ERP) peaking between 300 and 500ms after word onset over centro-parietal sites. The N400 amplitude is sensitive to contextual predictability, with less predictable words eliciting more negative amplitudes than highly predictable words(Kutas & Federmeier, 2011). Within the hierarchical predictive coding framework, the N400 is operationalized as a lexico-semantic prediction error signal, that emerges when meaning is inferred from an incoming speech signal(Fitz & Chang, 2019; Nour Eddine et al., 2024; Rabovsky & McRae, 2014). Following the predictive processing accounts of SSD and ASD, we expected distinct patterns of computational alterations and corresponding N400 modulations for autistic and schizotypal traits.

We hypothesized that individuals with increasing autistic traits would overweight sensory evidence, leading to heightened sensitivity to unexpected input, which should manifest in larger N400 differences between predictable and unpredictable sentences (**Figure 1A**). However, previous research on the N400 in ASD only partially supports this hypothesis as multiple studies report intact semantic processing(Braeutigam et al., 2008; Coderre et al., 2017; DiStefano et al., 2019; Kubinski et al., 2024; O’Rourke & Coderre, 2021; Pijnacker et al., 2010; Russo et al., 2012) or attenuated N400 effects, including children of different age groups(Cantiani et al., 2016; Dunn et al., 1999; Dunn & Bates, 2005; Manfredi et al., 2020; Márquez-García et al., 2022; McCleery et al., 2010; Ribeiro et al., 2013) and adults(Fishman et al., 2011; Kaplan-Kahn et al., 2021; Pijnacker et al., 2010; Ring et al., 2007).

In contrast, for increasing schizotypal traits we expected that individuals would demonstrate overweighting of high-level semantic prior beliefs, which may reduce sensitivity to unexpected input and should result in smaller N400 amplitude differences, indicating an attenuated prediction error signal (**Figure 1B**). Supporting this, previous studies using semantic priming tasks have repeatedly found a reduced N400 effect between semantically matching and mismatching word pairs across different disease stages, from subclinical schizotypal traits(Kostova et al., 2014; Prévost et al., 2011) to clinical high risk(Lepock et al., 2020, 2021a), and clinical stages(Mohammad & DeLisi, 2013; Wang et al., 2011).

Taken together, by investigating these effects in the general population, the present study aims to mechanistically test the ASD-SSD predictive processing spectrum hypothesis and provide a more nuanced computational and neurophysiological understanding of potential language processing deficits beyond clinical manifestations.

## METHODS

### PARTICIPANTS

Our sample consisted of 55 German native speakers (28 females, 27 males), aged between 18 and 35 (*M=*23.73, *SD=*3.85). All participants had normal hearing, and were right-handed, as assessed by a short form of Edinburgh Handedness Inventory(Veale, 2014). Prior to their participation, all participants completed a German 5-point Likert-scale version of the Schizotypal Personality Questionnaire (SPQ; (Raine, 1991; Wuthrich & Bates, 2005)) to assess schizotypal personality traits (*M*=86.56, *SD=*50.13), and the Autism Spectrum Quotient (AQ; (Baron-Cohen et al., 2001)) to measure subclinical autistic traits (*M*=21.09, *SD=*7.27). Additionally, we administered the subtests Vocabulary, Information, Similarities, and Comprehension of the Wechsler Adult Intelligence Scale IV (WAIS-IV;(Wechsler, 2012)) to obtain the verbal comprehension index (VCI) – a measure of verbal knowledge and reasoning abilities (*M=*110.56, *SD=*9.30). **Figure 2** shows the distribution of the subclinical and cognitive trait markers and their correlations. A subset of the participants (N=53) received ^1^H-MRS to assess levels of glutamate concentration in the ACC and the left DLPFC in a separate experimental session(Demler et al., 2023, 2024, 2025; Knolle et al., 2025). All participants gave informed written consent and received financial compensation or psychology course credits after completing the experiment. The study was approved by the Ethics Commission of the Technical University of Munich (approval number 2023/270 S-KH).

**Figure 2.**
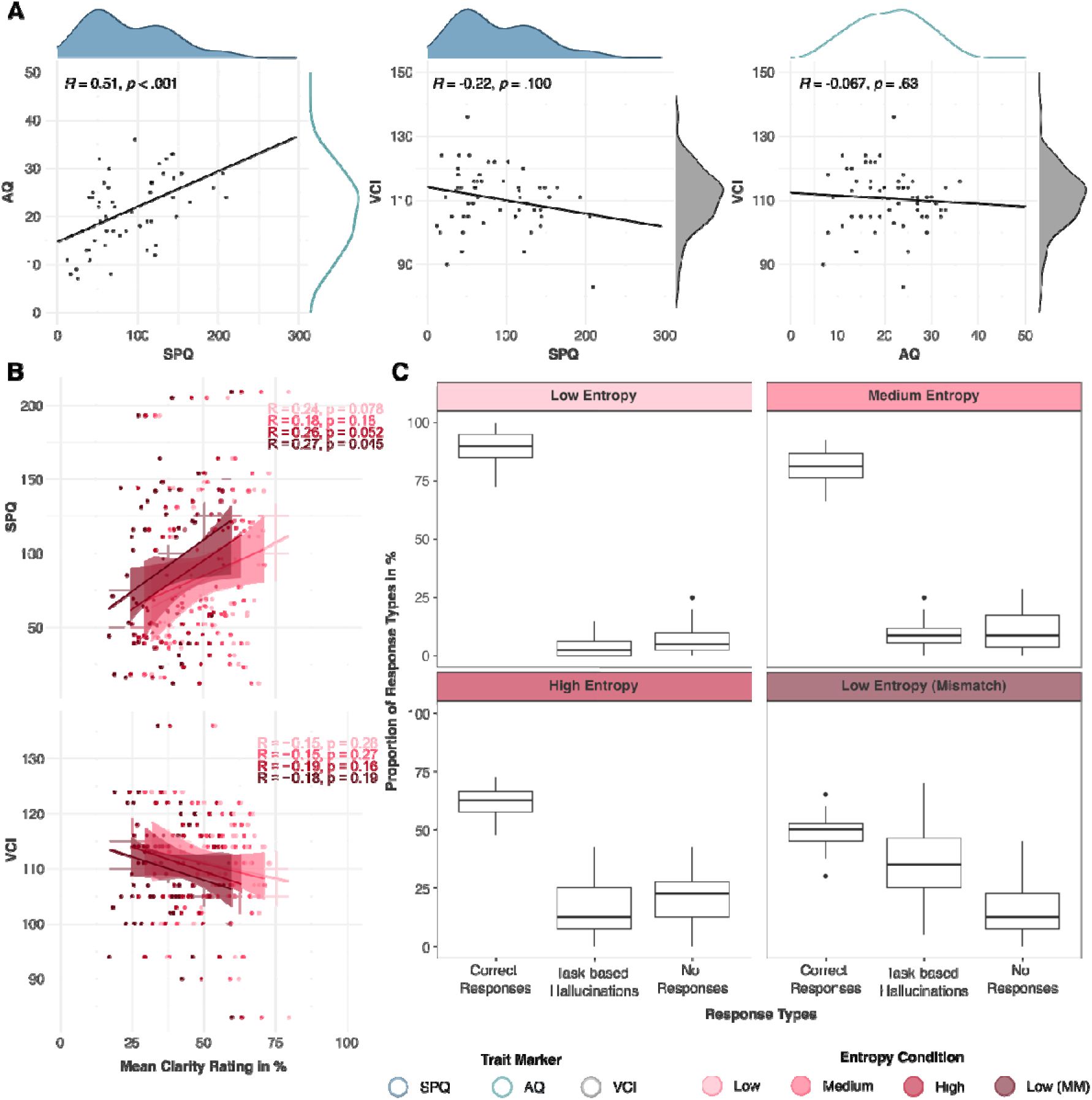
Subclinical, Cognitive Traits and Task Behavior. A) Association between subclinical and cognitive trait markers. Autistic and schizotypal traits were significantly correlated. Neither subclinical trait measure was significantly related to increased or decreased verbal comprehension ability. **B) Association between subclinical and schizotypal traits and subjective clarity ratings.** Higher levels of schizotypy showed a trend toward higher clarity ratings in low, medium and low entropy (mismatch conditions). Increasing verbal comprehension abilities showed a descriptively opposite, non-significant pattern. **C) Behavioral response types split by entropy conditions.** Correct responses decreased with decreasing predictability manipulations while task-based hallucinations increased. No responses stayed relatively stable.

### STIMULI AND DESIGN

The predictive language task was designed to investigate predictive language processing within the predictive processing framework (**Figure 1C**). The paradigm independently modulates the precision of high-level semantic prior beliefs and the precision of low-level sensory evidence. The semantic prior was formed by the presented sentence up to the sentence-final word. To modulate the semantic prior’s precision (i.e., the predictability), 40 high entropy sentences, 80 medium entropy sentences, and 80 low entropy sentences were selected for the experiment. Entropy was estimated using Shannon entropy(Shannon, 1948). For the high and medium entropy conditions, the word with the highest cloze probability was always selected as the sentence-final word. For the low entropy sentences, half of the sentences were completed by the word with the highest cloze probability, forming the low entropy condition, whereas the other half was completed by a syntactically plausible word with a cloze probability of 0 to maximize surprisal, forming the low entropy mismatch condition. Both Shannon entropy and cloze probability were assessed in an independent sample of participants(E. Sterner et al., 2025). The precision of the sensory evidence was modulated through different levels of noise vocoding on the sentence final word. Each word was degraded using 1, 3, 6 or 12 vocoder channels(Gaudrain & Başkent, 2015). Here, perceptual clarity is increasing with rising channel numbers(Cope et al., 2017). All sentence-final words were vocoded in all four levels of degradation. As each of the 200 sentences was only presented once, four experimental groups were formed to counterbalance clarity levels across participants and entropy conditions. After being assigned to an experimental group, stimulus presentation was fully randomized for every participant.

### TASK AND PROCEDURE

At the beginning of each trial, participants listened to a sentence beginning over headphones while focusing on a central fixation cross on the monitor. The degraded sentence-final word was presented after a delay period of 100ms. After listening to the complete sentence, participants were asked to rate the clarity of the sentence-final word by adjusting a slider bar between 0% (completely unclear) and 100% (completely clear). Participants were then instructed to say the word they understood, as soon as a microphone icon appeared in the center of the monitor. After their response, participants were asked to rate the confidence in their response by adjusting a slider bar between 0% (completely unsure) and 100% (completely sure). Afterwards, the next trial started. In total, participants completed 200 trials in four experimental blocks of 50 trials each.

Before the start of the experiment, participants completed seven practice trials with more detailed instructions to get familiar with the task. During the experiment, participants were seated centrally in front of a monitor (BenQ Corporation 24’’, resolution 1920x1080) in a darkened room and wore headphones (hama®, sensitivity 105 ± 3dB, frequency range 20Hz–20kHz, impedance 32W) with an integrated microphone (hama®, sensitivity -42 ±3dB, frequency range 50Hz–10kHz, impedance 2200 W, directivity omnidirectional) for stimulus presentation and recording. The experimental task was created using Psychopy3 Experiment Builder (v2022.1.1;(Peirce et al., 2019)) and presented in Psychopy Coder (v2022.1.1) which was synchronized with the recording of the EEG signals in BrainVision Recorder 1.24.0001 (Brain Products GmBH, Gilching, Germany).

### BEHAVIOURAL ANALYSIS

To investigate the effect of task manipulations and to replicate the results previously reported from a larger sample (Knolle et al., 2025), we performed two-factor repeated-measures ANOVAS on clarity and confidence ratings to examine the effects of predictability (i.e., entropy) and sensory evidence (i.e., channel degradation). Additionally, we performed repeated measures ANOVAs with entropy level as a factor to investigate the impact of sentence predictability on the distribution of behavioral response types (correct responses, no responses, task-based hallucinations). For all ANOVAs, Mauchly’s test was applied to assess sphericity; if violated, degrees of freedom were adjusted using Greenhouse-Geisser estimates. Significant main and interaction effects were further analyzed with Tukey post hoc tests. Associations with subclinical and cognitive trait markers were investigated using Pearson correlations.

### COMPUTATIONAL MODELLING

To mechanistically estimate how much participants relied on their semantic prior beliefs relative to the sensory evidence during our predictive language task, we applied a Bayesian Belief updating model. Specifically, we aimed to assess the relative weight of high-level semantic prior beliefs (i.e., entropy) in comparison to the sensory evidence (noise degradation, surprisal) and how this balance is modulated by individual differences in our trait (AQ/SPQ) and cognitive markers (VCI). The modelling approach assumes that participants aim to maximize the probability of providing a correct answer. In each trial, this probability takes a prior value after the presentation of the sentence up to the sentence-final word (semantic prior belief) and then undergoes an update to a posterior value after the presentation of the degraded sentence-final word (sensory evidence). This process follows a Bayesian update in a beta-Bernoulli model, where the observation follows a Bernoulli distribution as participant’s response either matches the presented sentence-final word (coded as 1) or does not (coded as 0). The prior and posterior distributions are modelled as beta distributions, reflecting uncertain probabilities of correctness.

According to Bayes’ rule, the expected correctness probability is updated using a precision-weighted prediction error:

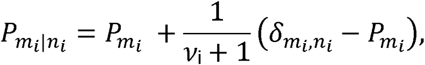

where *P_m_*_l_ and *P_m_*_l_*_|n_*_l_ represent the prior and posterior cloze probabilities, respectively; *m_i_* is the participant’s response in trial *i*, and *n_i_* is the corresponding sentence-final word. The Kronecker delta *o_m_*_l_*_,n_*_l_ indicates whether *m_i_ = n_i_* while *v*_i_ *>0* denotes the implied number of previous Bernoulli observations(Mathys & Weber, 2020), which can be directly interpreted as the weight of the prior relative to sensory evidence. The prediction in this model corresponds to the cloze probability of the participant’s response which is the only available information to the participant before hearing the sentence-final word. The prediction error corresponds to the difference of the correctness of the answer (1 or 0) minus the prediction and updates the expected correctness probability by adding it to the prediction after weighting by a factor of *1/(1 + v)*. *v* formalizes the weight of the prior relative to the sensory evidence and is an implied number of previous observations(Mathys & Weber, 2020). *v_i_* is modelled on a trial-by-trial basis within a hierarchical linear model that entails the task parameters entropy, cloze probability of the stimulus, and channel number. We further stratified all estimated by the participant’s subjective clarity rating to assess the direct effects. As we expected the prior weight to be modulated by participants’ trait markers, we included participant-level coefficient in the following hierarchical model for *log v_i_*:

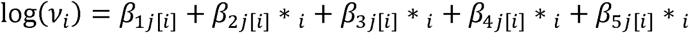

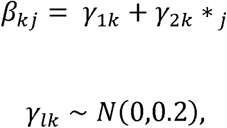

where *i* represents the trial, and *j*[*i*] the participant on trial *i*. *β_kj_* are participant-level coefficients, and *γ_lk_* are population-level coefficients. *v_i_* was estimated in logarithmic space as it is bounded below at zero in native space whereas in logarithmic space it is unbounded.

Hamiltonian Monte Carlo (HMC) sampling (four chains each with 2000 iterations including 1000 warm-up draws) with the rstan software package(Stan Development Team, 2018) was used to estimate the posterior distribution of all parameters (*γ_lk_*).

### EEG RECORDING AND PREPROCESSING

EEG was continuously recorded using a MR-compatible EEG 10-10 system with 64 Ag/AgCl ring electrodes (BrainVision Recorder, version 1.24.0001, Brain Products GmbH, Gilching, Germany) with a 1000Hz sampling rate and an online low pass filter of 250Hz. FCz was used as an active reference (online) and AFz as the ground. Impedance values were kept below 10kΩ throughout the experiment. EEG signals were amplified using a BrainAmp MRplus amplifier (BrainAmp, Brain Products GmbH, Gilching, Germany) in the range of ±3.28mV at a resolution of 0.1μV.

All preprocessing and analysis steps were performed in mne Python (1.5.0 (Larson et al., 2024)). During preprocessing, the ECG electrode was dropped for all participants due to noise. EEG data was filtered using a low-pass filter (cut-off frequency: 100Hz) and a high-pass filter (cut-off frequency: 0.1Hz). A notch filter was applied at 50Hz and the harmonics 100Hz and 150Hz. Automatic break detection was applied with a minimum break detection of 20s with a minimum gap of 2s after the previous break and before the next break. The EEG data was then rereferenced using a common average reference. An independent component analysis (ICA, picard algorithm, maximum number of iterations: 1000) was used for ocular artifact removal with Fp1 and F8 as “virtual” EOG channels. For the N400 ERP-analysis, the preprocessed EEG data was segmented into epochs from -200 to 1000ms around the target word onset for each of the four entropy conditions (low, medium, high, low mismatch). Only trials in which participants reported the correct sentence-final word were included. A baseline correction of the 200ms preceding the onset of the target word was applied. All epochs were automatically repaired or rejected based on local peak-to-peak amplitude, using the 0.4.1 autoreject package (Jas et al., 2017, 2018). The N400 was operationalized as the mean amplitude between 300-500ms after target word onset. For each participant, the mean N400 amplitudes were obtained by averaging epochs for each of the 15 centro-parietal electrodes (i.e., C3, C1, Cz, C2, C4, CP3, CP1, CPz, CP2, CP4, P3, P1, Pz, P2, P4) for each condition. Both the time frame and the selection of electrodes was based on previous literature in the field(Bohec et al., 2022; Del Goleto et al., 2016; Lepock et al., 2021b; Michaelov et al., 2024; Schoknecht et al., 2022).

### N400 ANALYSIS

Statistical analyses were performed in R Studio(R Core Team, 2020) and visualized using the ggplot2 package, version 3.5.1(Wickham, 2016), and the sjPlot package, version 2.8.17(Lüdecke, 2024). To investigate alterations in the mean N400 amplitude depending on the clinical scores and the four entropy conditions, we used linear mixed models (LMMs) performed with the lme4 package, version 1.1 – 35.5(Bates et al., 2015). The sentence’s entropy level (low, medium, high, low mismatch), the scaled subclinical (SPQ/AQ) and cognitive (VCI) trait markers, and the interaction between entropy condition and the scaled trait markers were included as fixed effects. The participant and the electrode position were defined as random intercept as we did not define hypotheses about spatial distribution or lateralization of effects. The LMMs were carried out separately for AQ, SPQ, and VCI. Tukey adjusted post hoc tests and simple slope analyses were conducted to follow up main effects and interaction effects using the emmeans package, version 1.10.4(Lenth, 2020), and the interactions package, version 1.2.0(Long, 2019).

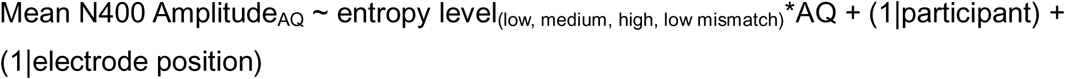

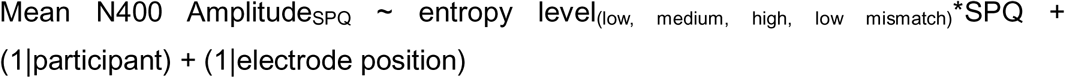

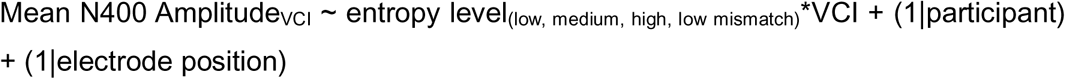

### P600 CONTROL ANALYSIS

Based on previous studies using related sentence processing paradigms (e.g., (Federmeier et al., 2007; Grisoni et al., 2021; Kutas & Federmeier, 2011; Michaelov et al., 2024)), we focused on the N400 as an electrophysiological signature of semantic prediction error processing. Visual inspection of the ERPs however suggested condition-related effects at later latencies. Given evidence that late positive components, such as the P600, can be sensitive to semantic processing (e.g., (Brothers et al., 2020; Delogu et al., 2019; Kuperberg et al., 2007, 2020)), we analyzed a later time window (600 to 900ms post final-word onset) using the same electrode cluster to assess whether condition- or trait-related effects extended beyond the N400 window. The same statistical approach was used as for the N400 analysis.

## RESULTS

### BEHAVIORAL ANALYSIS

To replicate the effect of task manipulations reported from a larger sample, we used repeated-measures ANOVAs. Subjective clarity and confidence ratings significantly decreased with increased degradation (fewer channels) and higher entropy (clarity: F(5.38,290.78)=52.37, p<0.001, general effect size=0.124; confidence: F(5.66,265.83)=42.30, p<0.001, effect size=0.143). Words with fewer channels and higher entropy were perceived as less clear, with lower confidence (**Tables S2 and S4**). These findings indicate that the subjective clarity rating not only reflects the sensory precision of the sentence-final word but is also an indirect measure of the precision of the semantic prior belief, as words following precise priors (lower entropy) were rated as clearer regardless of degradation level. Pearson correlation analyses indicated that increasing schizotypal were associated with increased clarity ratings whereas increasing verbal comprehension skills were associated with decreased clarity ratings, indirectly reflecting increased and decreased precision of the precision of semantic prior beliefs (**Figure S1**). Although these correlations did not reach significance, a similar effect size between schizotypy and clarity ratings was observed in a larger independent sample, suggesting that larger samples may be required to reliably detect these associations.

Misperceptions (task-based hallucinations) increased with higher entropy manipulations (F(1.89,102.05)=199.22, p<0.001, general effect size=0.617). Correct responses decreased (F(2.39,129.24)=553.00, p<0.001, general effect size=0.852) and non-responses increased (F(2.25,121.29)=64.98, p<0.001, general effect size=0.199) (**Fig.2, Tables S5, S6, S7**). This suggests increasing uncertainty, driven by semantic prior manipulations, contributes to task-based hallucinations. No significant correlations were observed between subclinical trait measures and behavioural performance across entropy conditions. However, verbal comprehension ability showed trend-level associations with performance, characterized by more correct responses in low- and medium-entropy conditions and fewer correct responses in high-entropy conditions, suggesting a potential interaction between verbal ability and contextual uncertainty.

### COMPUTATIONAL MODELLING

To investigate whether subclinical schizotypal and autistic traits are associated with an imbalance in the weighting of high-level semantic prior beliefs (i.e., entropy of the sentence beginning) and sensory evidence (i.e., noise degradation and surprisal of the sentence-final word), we performed a Bayesian belief updating model separately for each trait marker on the behavioral task data. A linear model including the trial-based task parameters (i.e., the sentence beginning’s entropy, sentence-final word’s cloze probability, the channel number with which the sentence-final word was presented, and the subjective clarity rating) and was used to estimate the strength *v* of the prior weight depending on the relative trait marker.

The model estimation converged and met all quality checks (**Figure S6 and S7**). **Table 1** and **Figure 4** show the parameter estimates using SPQ, AQ and VCI as model predictors. The modelling results showed that increasing SPQ induced a significant shift in the prior weight to the prior belief (coefficient gamma[2,1]), while AQ and VCI did not significantly alter the prior weight. Specifically, increasing SPQ by 10 points (range 0 -296) induced an increase of 0.39% in the weighting of semantic prior beliefs relative to the sensory evidence (i.e., exp(-0.30+0.19/50.13)/exp(-0.30)=1.0038, where 50.13 is SPQ SD, -0.30 is the gamma[1,1] intercept, and 0.19 is gamma[2,1], reflecting SPQ impact). Distribution analyses of the trait marker parameter gamma[2,1] using Tukey corrected Games-Howell post-hoc tests indicated that the effect of the trait markers on the prior weight was significantly different between all trait markers. The effect of SPQ was significantly stronger than the effect of AQ (0.15, 95% CI [0.15, 0.15], p<0.001). The effect of VCI was significantly weaker than the effect of SPQ (-0.34, 95% CI [-0.34, -0.33], p<0.001) and AQ (-0.19, 95% CI [-0.19, -0.18], p<0.001).

**Figure 4.**
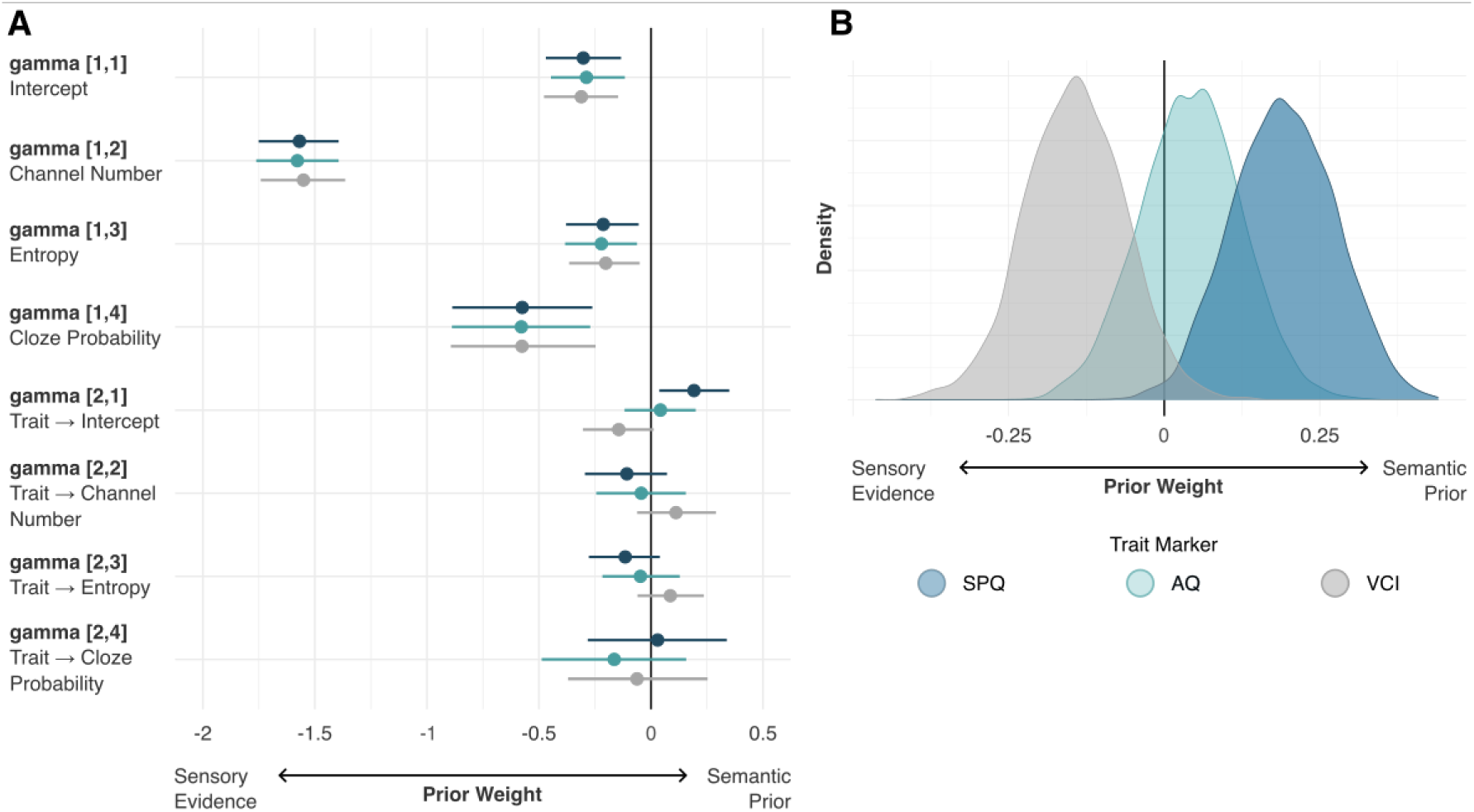
A) Parameter estimates of the linear model estimating the weight of the prior relative to the sensory evidence modulated by subject-level SPQ, AQ, and VCI scores. B) Differences in the distributions of the effect of SPQ, AQ, and VCI on the prior weight. SPQ: Schizotypal Personality Questionnaire(Raine, 1991; Wuthrich & Bates, 2005); AQ: Autism Spectrum Quotient(Baron-Cohen et al., 2001); VCI: Verbal Comprehension Index derived from the WAIS-IV(Wechsler, 2012)

**Table 1:**
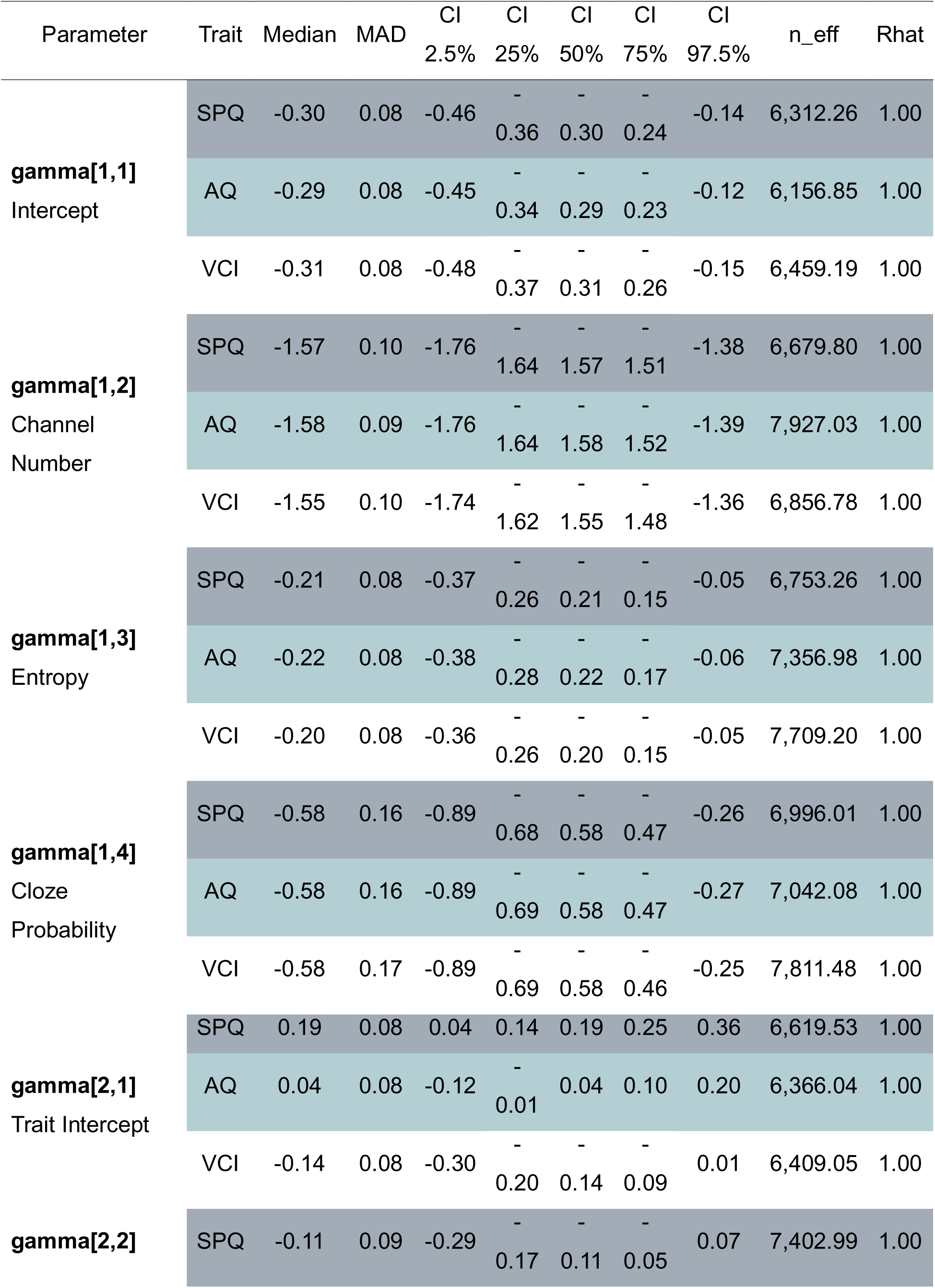
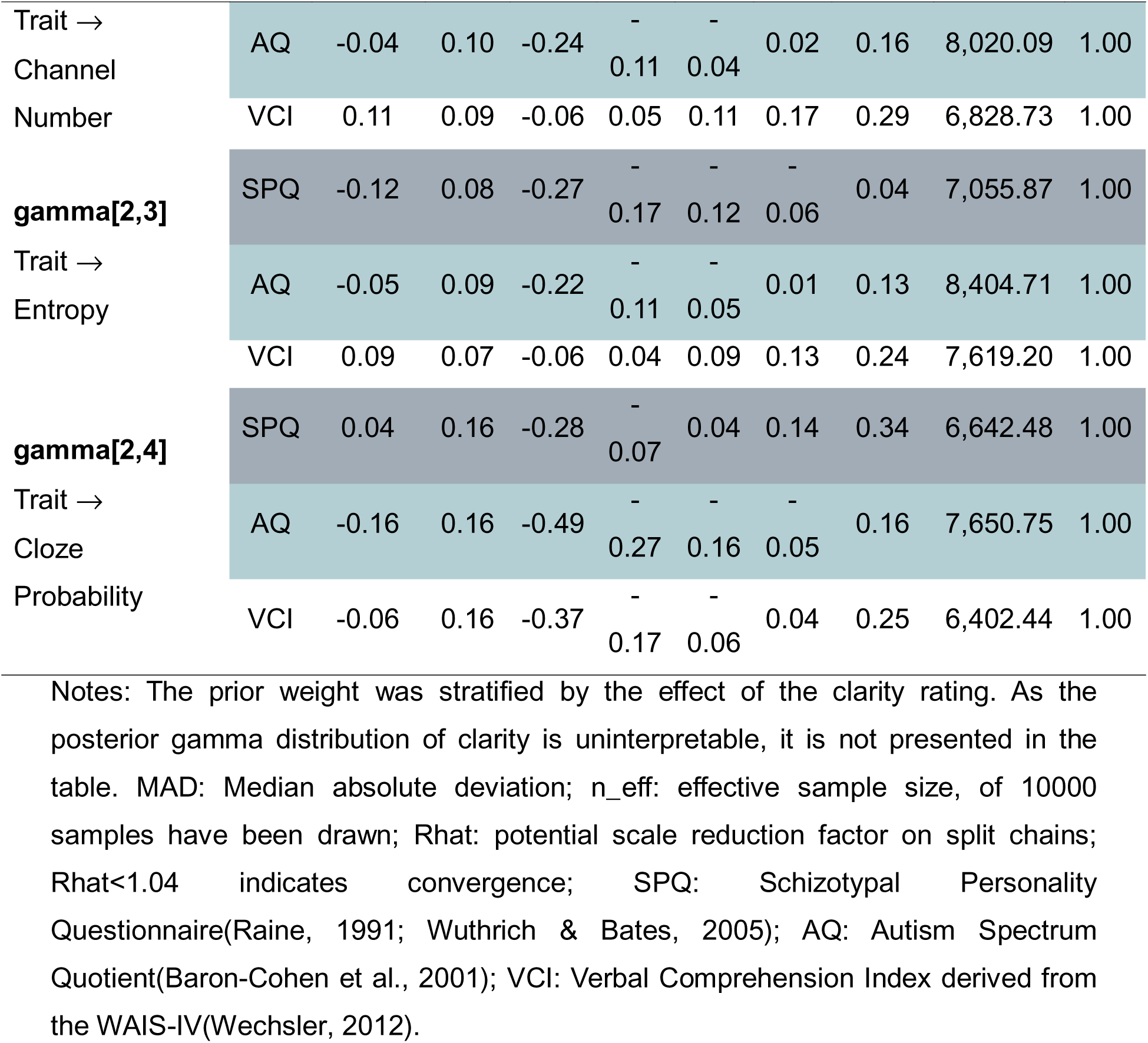
Parameter Estimates of Gammas Notes: The prior weight was stratified by the effect of the clarity rating. As the posterior gamma distribution of clarity is uninterpretable, it is not presented in the table. MAD: Median absolute deviation; n_eff: effective sample size, of 10000 samples have been drawn; Rhat: potential scale reduction factor on split chains; Rhat<1.04 indicates convergence; SPQ: Schizotypal Personality Questionnaire(Raine, 1991; Wuthrich & Bates, 2005); AQ: Autism Spectrum Quotient(Baron-Cohen et al., 2001); VCI: Verbal Comprehension Index derived from the WAIS-IV(Wechsler, 2012).

### N400 ANALYSIS

We performed linear mixed models (LMMs) separately for the subclinical and cognitive trait markers to investigate the effect of the trait markers and with sentence entropy levels on the mean N400 amplitude. Sentence entropy level (low, medium, high, low (mismatch)), subclinical (SPQ/AQ) and cognitive (VCI) trait markers, and their interactions were included as fixed effects, while the individual participant and 15 centro-parietal electrode positions (C3, C1, Cz, C2, C4, CP3, CP1, CPz, CP2, CP4, P3, P1, Pz, P2, P4) were included as random intercepts. The results are depicted in **Table 2** and **Figure 5**.

**Figure 5.**
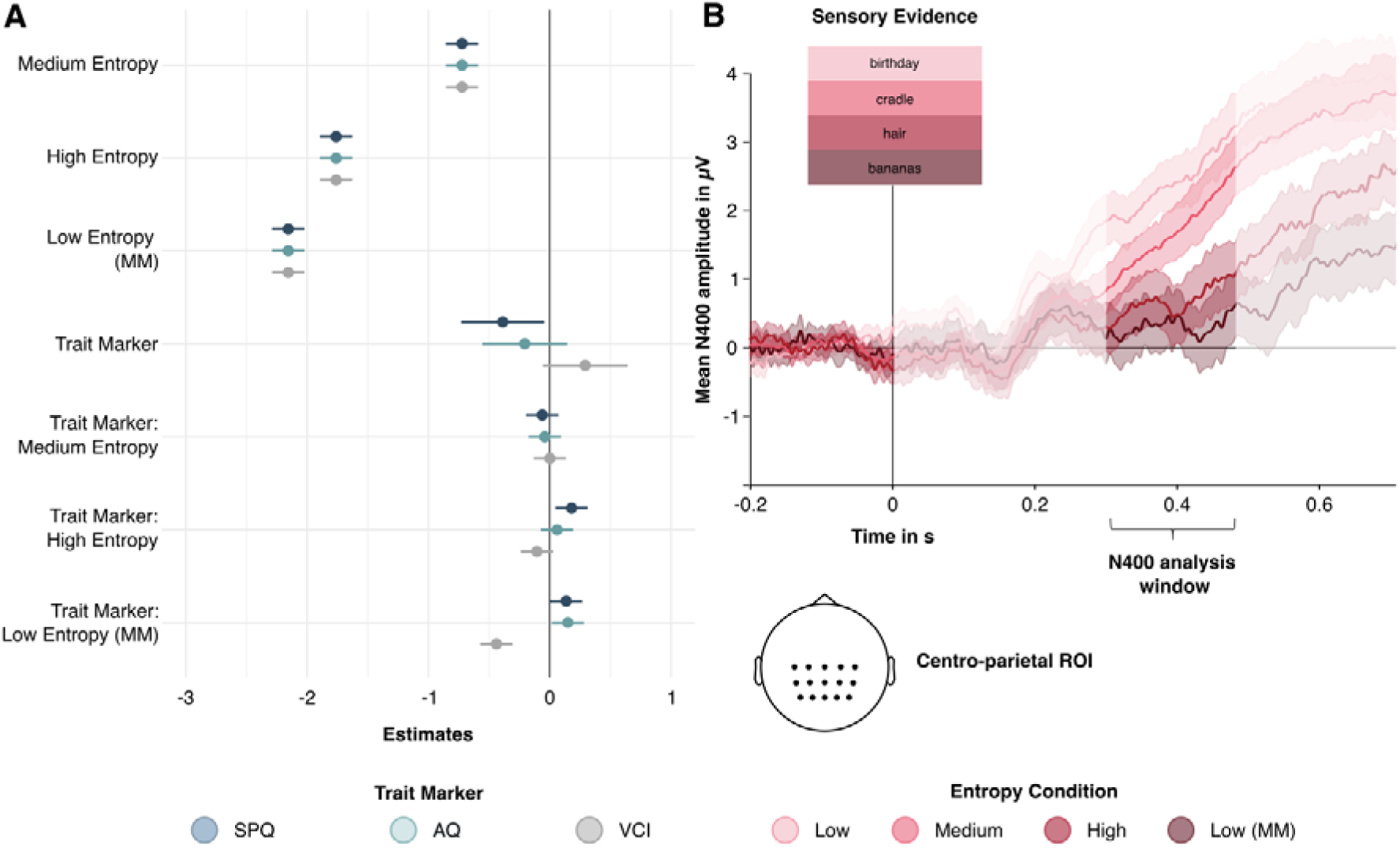
A) Linear mixed model results: Impact of subclinical (SPQ/AQ) and cognitive (VCI) trait markers and sentence entropy level on mean N400 amplitude. B) Mean N400 amplitude for sentence entropy levels time-locked to the onset of the sentence final word (e.g., low entropy: She wished her a happy <u>birthday</u>, medium entropy: The baby slept in the <u>cradle</u>, high entropy: In the bathtub swam a <u>hair</u>, low entropy (mismatch): He decorated the room with <u>bananas</u>; SPQ: Schizotypal Personality Questionnaire(Raine, 1991; Wuthrich & Bates, 2005); AQ: Autism Spectrum Quotient(Baron-Cohen et al., 2001); VCI: Verbal Comprehension Index derived from the WAIS-IV(Wechsler, 2012)

**Table 2:**
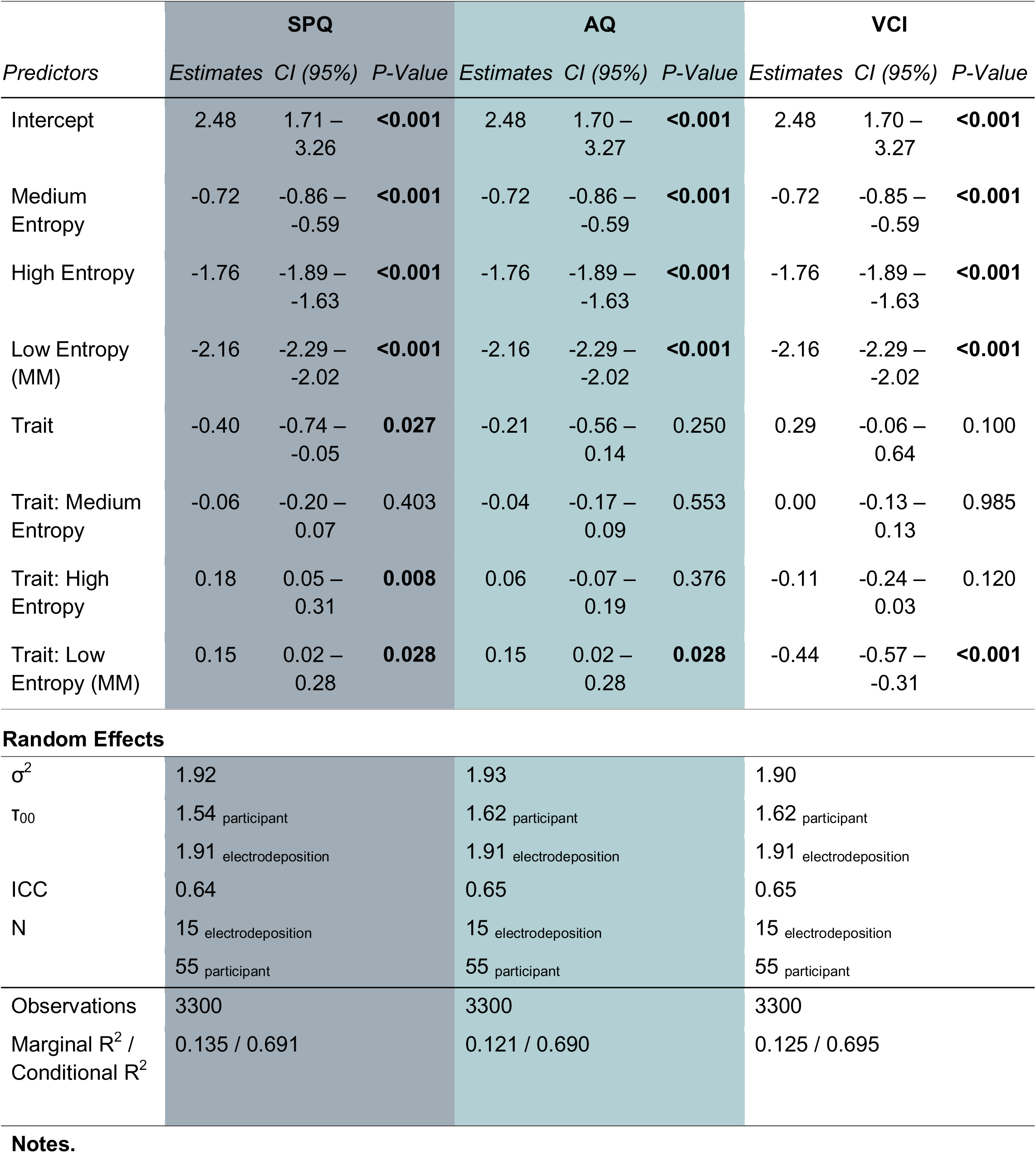
N400 LMM Results

| *Predictors* | **SPQ** |  |  | **AQ** |  |  | **VCI** |  |  |
| --- | --- | --- | --- | --- | --- | --- | --- | --- | --- |
|  | *Estimates* | *CI (95%)* | *P-Value* | *Estimates* | *CI (95%)* | *P-Value* | *Estimates* | *CI (95%)* | *P-Value* |
| Intercept | 2.48 | 1.71 – 3.26 | **<0.001** | 2.48 | 1.70 – 3.27 | **<0.001** | 2.48 | 1.70 – 3.27 | **<0.001** |
| Medium Entropy | -0.72 | -0.86 – -0.59 | **<0.001** | -0.72 | -0.86 – -0.59 | **<0.001** | -0.72 | -0.85 – -0.59 | **<0.001** |
| High Entropy | -1.76 | -1.89 – -1.63 | **<0.001** | -1.76 | -1.89 – -1.63 | **<0.001** | -1.76 | -1.89 – -1.63 | **<0.001** |
| Low Entropy (MM) | -2.16 | -2.29 – -2.02 | **<0.001** | -2.16 | -2.29 – -2.02 | **<0.001** | -2.16 | -2.29 – -2.02 | **<0.001** |
| Trait | -0.40 | -0.74 – -0.05 | **0.027** | -0.21 | -0.56 – 0.14 | 0.250 | 0.29 | -0.06 – 0.64 | 0.100 |
| Trait: Medium Entropy | -0.06 | -0.20 – 0.07 | 0.403 | -0.04 | -0.17 – 0.09 | 0.553 | 0.00 | -0.13 – 0.13 | 0.985 |
| Trait: High Entropy | 0.18 | 0.05 – 0.31 | **0.008** | 0.06 | -0.07 – 0.19 | 0.376 | -0.11 | -0.24 – 0.03 | 0.120 |
| Trait: Low Entropy (MM) | 0.15 | 0.02 – 0.28 | **0.028** | 0.15 | 0.02 – 0.28 | **0.028** | -0.44 | -0.57 – -0.31 | **<0.001** |

**Random Effects**
|  |  |  |  |  |  |
| --- | --- | --- | --- | --- | --- |
| $\sigma^2$ | 1.92 | | 1.93 | | 1.90 |
| $\tau_{00}$ | 1.54 participant | | 1.62 participant | | 1.62 participant |
|  | 1.91 electrodeposition |  | 1.91 electrodeposition |  | 1.91 electrodeposition |
| ICC | 0.64 |  | 0.65 |  | 0.65 |
| N | 15 electrodeposition |  | 15 electrodeposition |  | 15 electrodeposition |
|  | 55 participant |  | 55 participant |  | 55 participant |
| Observations | 3300 |  | 3300 |  | 3300 |
| Marginal R<sup>2</sup> / Conditional R<sup>2</sup> | 0.135 / 0.691 |  | 0.121 / 0.690 |  | 0.125 / 0.695 |

**Notes.**

For all three LMMs, there was a significant main effect of sentence entropy level. Paired Tukey corrected post-hoc comparison revealed a graded effect of sentence entropy level with a more negative N400 amplitude in the low entropy (mismatch) condition compared to the in the high entropy condition (SPQ/AQ/VCI: - 0.40, 95% CI=[-0.57, -0.22]); a more negative N400 amplitude in the high entropy condition compared to the medium entropy condition (SPQ/AQ/VCI: -1.04, 95% CI=[-1.21, -0.86]), and a more negative N400 amplitude in the medium entropy condition compared to the low entropy condition (SPQ/AQ/VCI:-0.72, 95% CI=[-0.90,-0.55]).

With increasing SPQ, there was a significant negative effect on the mean N400 amplitude in the low entropy baseline condition, indicating that the mean N400 amplitude is decreasing with increasing SPQ in the low entropy condition. Additionally, we found a significant interaction effect between SPQ and the high and low entropy (mismatch) condition compared to the effect of SPQ in the baseline condition, indicating that the mean N400 difference between the low entropy mismatch and the low entropy condition and the mean N400 amplitude difference between the high entropy and the low entropy condition decreased with increasing SPQ. Follow-up simple slope analyses confirmed that there was a significant decrease of the mean N400 amplitude with increasing SPQ in the low entropy condition (-0.40, SE=0.17, t= -2.28, *p*=0.027). In addition, the mean N400 amplitude also significantly decreased in the medium entropy condition (-0.45, SE=0.17, t= - 2.60, *p*=0.012). There was no significant change in the high (-0.22, SE=0.17, t=-1.24, *p*=0.221) and low entropy mismatch (-0.25, SE=0.17, t= -1.42, *p*=0.162) condition with increasing SPQ on their own.

For the LMM including AQ, there was a significant interaction effect between AQ and the low entropy (mismatch) condition compared to the effect of AQ in the baseline condition, suggesting that the mean N400 difference between the low entropy and low entropy mismatch condition decreased with increasing AQ. Follow-up simple slope analyses did not reveal a significant change of the mean N400 amplitude with increasing AQ in any condition on their own (low entropy: -0.21, SE=0.18, t= -1.15, *p*=0.254; medium entropy: -0.25, SE=0.18, t= -1.38, *p*=0.173; high entropy: -0.15, SE=0.18, t= -0.81, *p*=0.420; low entropy (mismatch): -0.06, SE=0.18, t= -0.31, *p*=0.757).

For the LMM including VCI, there was a significant interaction effect between VCI and the low entropy (mismatch) condition compared to the effect of VCI in the baseline condition, indicating an increasing mean N400 difference between the low entropy and low entropy mismatch condition with increasing VCI. Follow-up simple slope analyses did not reveal a significant change of the mean N400 amplitude with increasing VCI in any condition on their own (low entropy: 0.29, SE=0.18, t= 1.64, *p*=0.106; medium entropy: 0.295, SE=0.18, t=1.65, *p*=0.104; high entropy: 0.188, SE=0.18, t=1.05, *p*=0.297; low entropy (mismatch): -0.145, SE=0.18, t=-0.82, *p*=0.419).

### P600 CONTROL ANALYSIS

After visual inspection of the ERPs, we performed linear mixed models (LMMs) separately for the subclinical and cognitive trait markers to investigate the effect of the trait markers and with sentence entropy levels during a later time frame (600 to 900ms post final-word onset). Sentence entropy level (low, medium, high, low (mismatch)), subclinical (SPQ/AQ) and cognitive (VCI) trait markers, and their interactions were included as fixed effects, while the individual participant and 15 centro-parietal electrode positions (C3, C1, Cz, C2, C4, CP3, CP1, CPz, CP2, CP4, P3, P1, Pz, P2, P4) were included as random intercepts. The results are depicted in **Table S8**.

For all three LMMs, there was a significant main effect of sentence entropy level. Paired Tukey corrected post-hoc comparison revealed a similar graded effect of sentence entropy level as in the N400 analyses with a more negative amplitude in the low entropy (mismatch) condition compared to the in the high entropy condition (SPQ/AQ/VCI: -0.99, 95% CI=[-1.21, -0.76]); a more negative N400 amplitude in the high entropy condition compared to the medium entropy condition (SPQ/AQ/VCI: - 0.95, 95% CI=[-1.17, -0.72]). In contrast to the results in the N400 time-window, there was no significant amplitude difference between the medium entropy condition and the low entropy condition (SPQ/AQ/VCI:-0.05, 95% CI=[-0.28, 0.18]).

In contrast to the N400 time-window, increasing SPQ did not have a significant effect in the low entropy baseline condition. We found a significant interaction effect between SPQ and the medium and low entropy (mismatch) condition compared to the effect of SPQ in the baseline condition, indicating that the mean amplitude difference between the medium entropy and the low entropy condition and the mean amplitude difference between the low entropy mismatch condition and the low entropy condition increased with increasing SPQ. Follow-up simple slope analyses did not reveal a significant change of the mean amplitude with increasing SPQ in any condition on their own (low entropy: 0.02, SE=0.23, t=0.09, *p*=0.933; medium entropy=-0.21, SE=0.23, t=-0.92, *p*=0.359; high entropy=-0.09, SE=0.23, t=-0.39, *p*=0.698; low entropy (mismatch): -0.33, SE=0.23, t=-1.43, *p*=0.157).

For the LMM including AQ, there was a significant interaction effect between AQ and the low entropy (mismatch) condition compared to the effect of AQ in the baseline condition, suggesting that the mean amplitude difference between the low entropy and low entropy mismatch condition increased with increasing AQ. Follow-up simple slope analyses did not reveal a significant change of the mean amplitude with increasing AQ in any condition on their own (low entropy: 0.05, SE=0.23, t=0.23, *p*=0.819; medium entropy=0.09, SE=0.23, t=0.39, *p*=0.696; high entropy=0.20, SE=0.23, t=0.57, *p*=.572; low entropy (mismatch): 0.00, SE=0.23, t=0.00, *p*=0.999).

For the LMM including VCI, there was a significant interaction effect between VCI and the low entropy (mismatch) condition compared to the effect of VCI in the baseline condition, indicating an increasing mean amplitude difference between the low entropy and low entropy mismatch condition with increasing VCI. Follow-up simple slope analyses did not reveal a significant change of the mean N400 amplitude with increasing VCI in any condition on its own (low entropy: 0.25, SE=0.23, t=1.12, *p*=0.267; medium entropy=0.41, SE=0.23, t=1.80, *p*=0.077; high entropy=0.27, SE=0.23, t=1.21, *p*=0.230; low entropy (mismatch): -0.17, SE=0.23, t=-0.51, *p*=0.610).

## DISCUSSION

This study aimed to improve the computational and neurophysiological understanding of language processing deficits in subclinical expressions of ASD and SSD within the framework of predictive processing. We used an ecologically valid auditory comprehension task designed to directly manipulate the precision of high-level semantic prior beliefs and sensory evidence and applied hierarchical Bayesian belief updating modeling and EEG, focusing on the processing of semantic precision-weighted prediction errors as indexed by the N400 component. Computational modeling revealed that increasing schizotypal traits were associated with a significant overweighting of prior beliefs, while autistic traits did not show a significant shift. Linear mixed models on the mean N400 amplitudes further indicated that this schizotypy-related overweighting of semantic prior beliefs was reflected in a reduced semantic prediction error signal, indexed by smaller N400 differences between low entropy sentences and both high and low-mismatch sentences. A similar pattern emerged for increasing autistic traits, though the effect was weaker and less distinct as the N400 difference between low and low mismatch sentences also decreased for increasing autistic traits.

Consistent with the ASD-SSD predictive processing spectrum hypothesis, our computational results showed that increasing schizotypy was related to an overweighting of high-level semantic prior beliefs. This finding aligns with both computational studies(Cassidy et al., 2018; Haarsma et al., 2020; Knolle et al., 2025; A. R. Powers et al., 2017; Stuke et al., 2021; Teufel et al., 2015), which found stronger prior weighting across different cognitive and perceptual domains, as well as studies using sine-wave speech stimuli, which suggest overly strong priors for perceiving speech in noise in SSDs(Alderson-Day et al., 2017, 2022; Castiello et al., 2025; Kafadar et al., 2022). At the neurophysiological level, this shift was manifested in a reduced prediction error signal, indexed by an attenuated mean N400 amplitude difference between low entropy and both low-entropy mismatch and high-entropy sentences. This aligns with previous findings from semantic priming paradigms indicating reduced N400 effect across illness stages(Kostova et al., 2014; Lepock et al., 2020, 2021b; Mohammad & DeLisi, 2013; Prévost et al., 2011; Wang et al., 2011). By using an auditory sentence processing design, our findings adds to the limited number of studies which have moved beyond word-pair designs(Bohec et al., 2022; Del Goleto et al., 2016; Pinheiro et al., 2013; Salisbury, 2010), demonstrating that impaired use of semantic context also affects higher-level auditory semantic processing. Notably, we found that the reduced N400 effect with increasing schizotypy was driven specifically by impaired processing of predictable input as N400 amplitudes became more negative for predictable contexts (low and medium entropy sentences) with schizotypy increasing. This is also in line with previous semantic priming literature and sentence processing studies which identified more negative amplitudes for related compared to unrelated word pairs as the source of a diminished N400 effects(Del Goleto et al., 2016; Kiang et al., 2014; Lepock et al., 2019; Pinheiro et al., 2015; Salisbury, 2008). By introducing multiple levels of contextual predictability, our results extend this literature by showing that these alterations are not restricted to highly predictable associations but also occur in sentences that provide only moderate, yet meaningful, predictive cues.

In contrast to predictive processing accounts of ASD and the proposed ASD-SSD predictive processing continuum(Haker et al., 2016; Lawson et al., 2014; Pellicano & Burr, 2012; van Schalkwyk et al., 2017), we did not observe an overweighting of sensory evidence with increasing autistic traits. Most empirical support for such overweighting comes from visual perceptual(Amoruso et al., 2019; Eckert et al., 2024; Karvelis et al., 2018; Lieder et al., 2019; Tarasi et al., 2023; Zaidel et al., 2015) and social decision-making(Balsters et al., 2017; Ganglmayer et al., 2020; Kinard et al., 2020; Sevgi et al., 2020) studies, and the overall evidence is highly heterogeneous (see(Angeletos Chrysaitis & Seriès, 2023) for a review). Given these mixed findings, it may be possible that predictive processing impairments in ASD are domain-specific, with language processing remaining intact. This, however, would be difficult to reconcile with the clinical observation that that individuals with ASD consistently show robust impairments in speech processing(Groen et al., 2008; Tager-Flusberg et al., 2005).

Additionally, our findings argue against this domain-specific explanation as we observed changes in prediction error processing on an electrophysiological level. However, instead of larger N400 effects which would reflect sensory overweighting, we observed attenuated prediction error signaling as indexed by smaller mean N400 amplitude difference between low-entropy and low-entropy mismatch sentences. This result is consistent with previous studies reporting reduced, rather than enhanced, N400 effects in ASD(Cantiani et al., 2016; Dunn et al., 1999; Dunn & Bates, 2005; Fishman et al., 2011; Kaplan-Kahn et al., 2021; Manfredi et al., 2020; Márquez-García et al., 2022; McCleery et al., 2010; Pijnacker et al., 2010; Ribeiro et al., 2013; Ring et al., 2007), although the evidence is more heterogeneous than for SSD(Braeutigam et al., 2008; Coderre et al., 2017; DiStefano et al., 2019; Kubinski et al., 2024; O’Rourke & Coderre, 2021; Pijnacker et al., 2010; Russo et al., 2012).

Taken together, while computational modeling did not reveal altered weighting with increasing autistic traits, the neurophysiological results instead suggest a subtle overweighting of prior beliefs, similar to the pattern seen with increasing schizotypy. Given that the observed N400 alterations were weaker for increasing autistic traits, and that the N400 analysis averaged across entropy levels, such prior overweighting may emerge only under specific conditions not captured by task-level estimates. Future single-trial analyses which integrate computational modeling directly with EEG analyses may therefore provide more fine-grained insights.

Taken together, our results challenge the hypothesis that subclinical expressions of ASD and SSD exhibit diametric alterations in predictive language processing, as would be expected from an ASD-SSD predictive processing continuum. Instead, both autistic and schizotypal traits were associated with attenuated N400 prediction error responses, suggesting a shared tendency to overweight semantic prior beliefs rather than the predicted opposing computational pattern. In a recent study(E. F. Sterner et al., 2025), we also found that individuals with increasing schizotypal and autistic traits exhibit reduced temporal stability of semantic predictions, particularly in highly predictable (i.e., low entropy) contexts in which stable predictions typically facilitate processing. Together with the present findings, these results point to a similar impairment in both subclinical spectra to use contextual information to efficiently guide language comprehension.

This interpretation aligns with psychometric evidence indicating that ASD and SSD(Chisholm et al., 2015; Jutla et al., 2022; Romanovsky et al., 2025; Schalbroeck et al., 2023) share substantial overlap in specific symptom domains which is also mirrored in the relationship between autistic and schizotypal personality traits(Nenadić et al., 2021; Zhou et al., 2019), such as apathy, reduced affect sharing, and social withdrawal. According to these large-scale studies, psychotic symptoms (i.e., hallucinations and delusions) may more clearly differentiate the two spectra which may explain why studies using perceptual paradigms identify diametrical alterations in predictive processing mechanisms. At the same time, however, other studies demonstrate significant psychotic vulnerability in ASD, with higher rates psychotic experiences and psychotic disorders compared to the general population(Kiyono et al., 2020; Lai et al., 2019; Lugo-Marín et al., 2019). This indicates that the relationship between ASD and SSD is more complex than a single linear (predictive processing) continuum can capture. Related to this, previous empirical research in SSD point to multiple predictive processing alterations coexisting on different processing levels, such as low-level sensory overweighting resulting in amplified prediction error signaling, alongside overweighting of high-level priors. These distinct mechanisms may map onto different symptoms and may rely on different neurobiological mechanisms(Corlett et al., 2010; Goodwin et al., 2025; Sterzer et al., 2018). A similarly domain-specific framework may explain the heterogeneous and often inconsistent evidence for sensory overweighting in ASD and could potentially offer a deeper understanding of the computational phenotype.

As a cognitive trait measure we assessed the verbal comprehension index (VCI) of the WAIS-IV(Wechsler, 2012) to investigate whether verbal knowledge and reasoning abilities influence the weighting of semantic prior beliefs and sensory information. Although computational modeling did not reveal a significant shift in the weighting there was a trend toward greater reliance on sensory evidence with increasing verbal ability scores (MD=-0.14; 95% CI=[-0.30; 0.01]). Comparisons of posterior gamma distributions support this trend, as the increasing verbal abilities significantly shifted the weighting parameter to negative values in comparison to the effect of autistic and schizotypal personality traits. Electrophysiologically, this was reflected in an enhanced difference between low entropy and low entropy mismatch conditions, indicating a larger prediction error response. This aligns with previous studies showing that the N400 is not only influenced by global task manipulations (e.g., word expectancy) but also by individual differences in statistical learning (Munding et al., 2025), working-memory performance (e.g., Kim et al., 2018; Nakano et al., 2010), and language skills (e.g., Milligan et al., 2025), with higher skills yielding larger N400 effects. Our computational modelling results indicate a mechanistic account for this observation as higher verbal abilities enable stronger emphasis on the incoming sensory information. A reason for this might be that verbal abilities may help shaping a most efficient predictive framework with reliable semantic predictions, allowing more capacity to process sensory information and integrating them into the semantic predictive model. In case of unexpected incoming sensory information when semantic predictions are violated, as reflected in the difference between low entropy and low entropy mismatch sentences, the resulting prediction error would consequently be larger. Importantly, the opposing directions of the weighting effects for schizotypy inducing prior overweighting and verbal abilities inducing sensory overweighting were mirrored in opposing N400 effects. This suggests that our weighting parameter successfully captures how these distinct mechanisms differentially shape electrophysiological responses.

Notably, there was no significant association between schizotypy and verbal comprehension skills, indicating that the overweighting of prior beliefs in schizotypy is not attributable to reduced verbal reasoning abilities.

### LIMITATIONS, CONTROL ANALYSES AND FUTURE DIRECTIONS

In the present study, we focused on the N400 ERP component, as previous linguistic work using sentence-processing paradigms has linked semantic prediction error processing to this time window(Fitz & Chang, 2019; Nour Eddine et al., 2024; Rabovsky & McRae, 2014). Visual inspection of our data additionally suggested effects at later latencies, potentially indicative of modulations in the P600 or other late positive components. Although the P600 has been classically associated with syntactic violations or syntactically complex sentences (Hagoort et al., 1993; Kaan et al., 2000; Osterhout & Holcomb, 1992), accumulating evidence shows that it is also sensitive to semantic processing, with semantic violations eliciting enhanced positive amplitudes related to higher-level semantic integration processes (e.g., Brothers et al., 2020; Delogu et al., 2019; Kuperberg et al., 2007, 2020). Focusing on late positive components, previous studies have also identified alterations particularly in SSD und subclinical cohorts using different paradigms(Del Goleto et al., 2016; Kuperberg et al., 2006, 2018; Lee et al., 2016; Ruchsow et al., 2003). In the subclinical sample of the present study, however, control analyses indicated that the modulation observed during the N400 window extended into later time periods, with low entropy sentences eliciting more positive mean amplitudes than high-entropy and low-entropy mismatch sentences. This pattern is inconsistent with previous P600 findings, which would predict increased positivity for semantic unpredictability or violations. One possibility is that the robust N400 effect overlapped with or masked later positive components. Alternatively, the absence of a distinct P600 may relate to paradigm differences as previous studies often employed highly controlled designs with context built across multiple sentences which may require stronger semantic integration compared to shorter contexts (Brothers et al., 2020; Del Goleto et al., 2016; Delogu et al., 2019; Kuperberg et al., 2020). More fine-grained analyses of sentence types, time windows, and spatial distributions may help disentangle early semantic prediction error effects from later integrative processes but this will require further paradigm optimization in future research.

A strength of the experimental paradigm was the independent manipulation of the precision of semantic prior beliefs through sentence entropy and sensory precision through auditory degradation and contextual surprisal which is crucial for assessing the relative contributions of prior beliefs and sensory evidence to potential weighting imbalances though computational modelling. For the ERP analysis, trials were averaged as a function of sentence entropy for correct responses in line with previous studies, collapsing across auditory degradation levels. This approach increased signal-to-noise ratio for the components of interest but limited the ability to examine effects of sensory precision in isolation at the ERP level due to low trial counts per condition. Importantly, auditory degradation levels were counterbalanced across sentences and participants, reducing the likelihood that the observed effects were driven by systematic sensory differences. Future analyses combining single-trial ERP analyses with computational parameters which integrate the different task manipulations may allow a more fine-grained investigation of how sensory and prior precision jointly shape neural responses during language processing.

Finally, future studies are needed to investigate these mechanisms in clinical cohorts since the present study was conducted in a healthy subclinical sample. As both ASD and SSD show large heterogeneity in their cognitive symptom profile, future research would also benefit from larger sample sizes to stratify individuals better. Whole-brain modeling approaches and advanced single-trial analyses will additionally help to directly map computational parameters and electrophysiological signatures.

## Data Availability

All data produced in the present study are available upon reasonable request to the authors

