## Supplementary Material for "Altered Semantic Prediction Error Processing with Increasing Schizotypal and Autistic Traits"

**Supplementary Results**

**Statistical results showing the impact sentence entropy and target word degradation on clarity and confidence rating**

Clarity rating

We performed a two-factor repeated measure ANOVA to examine the effect of the entropy channel manipulation on mean subjective confidence ratings. Mauchly’s test indicated that the assumption of sphericity had been violated for the main effects of condition (W = 0.63, p < .001, ε = .65) and channel number (W = 0.67, p < .05, ε = .70) and for the interaction effect of condition and channel number (W = 0.60, p < .001, ε = .67). Consequently, degrees of freedom were corrected using Greenhouse-Geisser estimates of sphericity. All effects reported are significant at p < .05. There was a significant main effect of entropy condition, a main effect of channel number as well as an interaction effect of condition and channel number on the mean clarity rating (see **Table S1**). Tukey post hoc tests of the significant interaction effect revealed that for channel number 12, 6, and 3, the mean clarity rating significantly decreased from low entropy to medium entropy, to high entropy sentences, with the target word of the low entropy mismatch condition being perceived as the most unclear (see **Table S2**). For the 12 channel condition, the target word of a low and medium entropy sentence was perceived as clearer than during the high entropy or low entropy mismatch condition, however there was no significant difference between the low and medium entropy condition. For the 1 channel condition, the target word of a low entropy sentence was perceived as clearer than during the high entropy or low entropy mismatch condition, however there was no significant difference between the medium entropy condition, and between the medium and low entropy mismatch condition.

Pearson correlation analysis between subjective clarity ratings and subclinical and cognitive trait markers are depicted in **Figure S1**.

| **Table S1: Results of the repeated measures ANOVA investigating the effect of condition and channel number on the mean clarity rating** | | | | | |
| --- | --- | --- | --- | --- | --- |
| **Effect** | **df** | **MSE** | **F** | **ges** | **p-value** |
| condition | 1.88, 101.67 | 109.14 | 285.31 *** | .279 | <.001 |
| channel_nr | 2.02, 108.93 | 282.50 | 1134.91 *** | .810 | <.001 |
| condition:channel_nr | 5.38, 290.78 | 75.86 | 52.37 *** | .124 | <.001 |

| **Table S2: Results of Tukey post hoc tests investigating the interaction effect of entropy condition and channel number on the subjective mean clarity rating** | | | | | | |
| --- | --- | --- | --- | --- | --- | --- |
| contrast | channel_nr | estimate | SE | df | t.ratio | p.value |
| el - em | 12 | 0.67 | 0.66 | 54 | 1.01 | 0.74 |
| el - eh | 12 | 4.16 | 0.93 | 54 | 4.45 | 0.00 |
| el - elmm | 12 | 19.45 | 1.86 | 54 | 10.43 | 0.00 |
| em - eh | 12 | 3.49 | 0.96 | 54 | 3.64 | 0.00 |
| em - elmm | 12 | 18.78 | 1.82 | 54 | 10.31 | 0.00 |
| eh - elmm | 12 | 15.29 | 2.03 | 54 | 7.55 | 0.00 |
| el - em | 6 | 3.03 | 0.81 | 54 | 3.72 | 0.00 |
| el - eh | 6 | 13.66 | 1.51 | 54 | 9.02 | 0.00 |
| el - elmm | 6 | 30.78 | 1.67 | 54 | 18.42 | 0.00 |
| em - eh | 6 | 10.63 | 1.32 | 54 | 8.07 | 0.00 |
| em - elmm | 6 | 27.75 | 1.43 | 54 | 19.39 | 0.00 |
| eh - elmm | 6 | 17.11 | 1.63 | 54 | 10.53 | 0.00 |
| el - em | 3 | 5.23 | 1.34 | 54 | 3.91 | 0.00 |
| el - eh | 3 | 22.63 | 1.51 | 54 | 15.01 | 0.00 |
| el - elmm | 3 | 30.45 | 1.95 | 54 | 15.58 | 0.00 |
| em - eh | 3 | 17.40 | 1.68 | 54 | 10.35 | 0.00 |
| em - elmm | 3 | 25.22 | 2.03 | 54 | 12.44 | 0.00 |
| eh - elmm | 3 | 7.82 | 1.52 | 54 | 5.13 | 0.00 |
| el - em | 1 | 1.30 | 0.58 | 54 | 2.25 | 0.12 |
| el - eh | 1 | 4.25 | 0.63 | 54 | 6.71 | 0.00 |
| el - elmm | 1 | 2.62 | 0.77 | 54 | 3.39 | 0.01 |
| em - eh | 1 | 2.94 | 0.50 | 54 | 5.85 | 0.00 |
| em - elmm | 1 | 1.32 | 0.59 | 54 | 2.24 | 0.13 |
| eh - elmm | 1 | -1.63 | 0.53 | 54 | -3.09 | 0.02 |

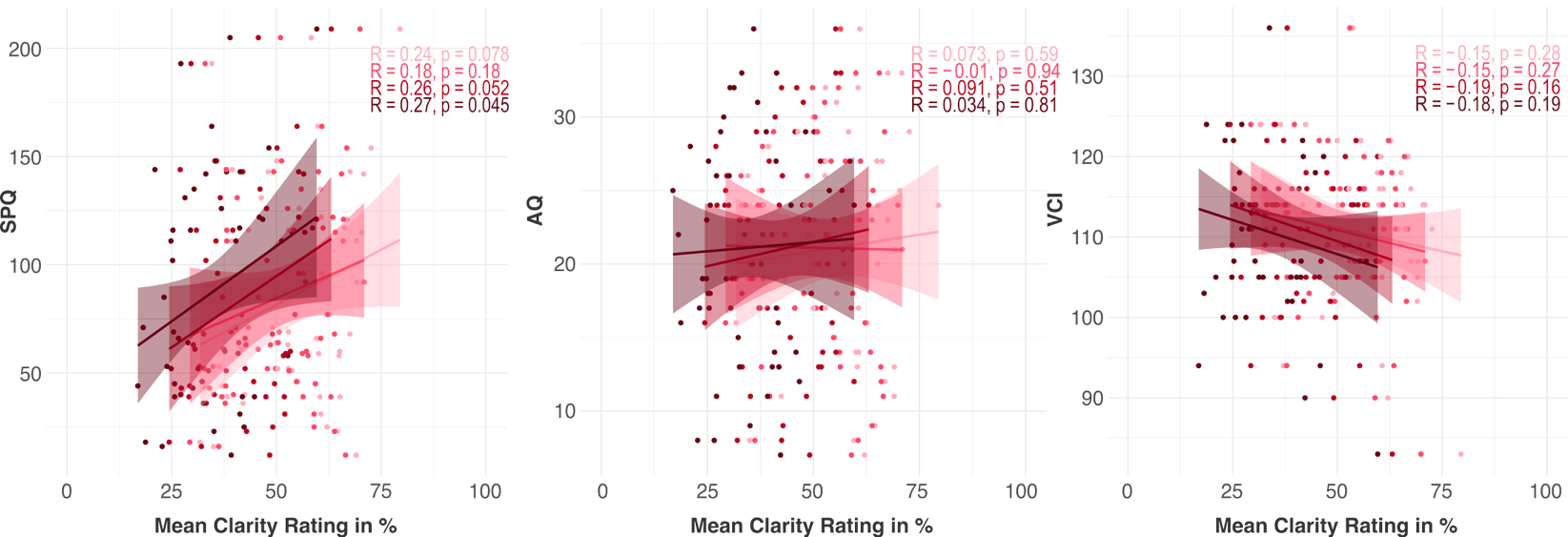

**Figure S1.** Scatter plots between subclinical and cognitive trait markers and subjective clarity ratings. P-Values are uncorrected.

R: Pearson Correlation Coefficient; SPQ: Schizotypal Personality Questionnaire(Raine, 1991; Wuthrich & Bates, 2005); AQ: Autism Spectrum Quotient(Baron-Cohen et al., 2001); VCI: Verbal Comprehension Index derived from the WAIS-IV(Wechsler, 2012)

Confidence rating

We performed a two-factor repeated measure ANOVA to examine the effect of the entropy channel manipulation on mean subjective confidence ratings. Mauchly’s test indicated that the assumption of sphericity had been violated for the main effects of condition (W = 0.39, p < .001, ε = .64) and channel number (W = 0.17, p < .001, ε = .49) and for the interaction effect of condition and channel number (W = 0.03, p < .001, ε = .63). Consequently, degrees of freedom were corrected using Greenhouse-Geisser estimates of sphericity. All effects reported are significant at p < .05. There was a significant main effect of entropy condition, channel number as well as a significant interaction effect of condition and channel on the mean confidence rating (see **Table S3**). Tukey post hoc tests of the significant interaction indicated that for the channel numbers 12, 6, and 3, the mean confidence ratings significantly decreased from low entropy to medium entropy to high entropy sentences, with responses in the low entropy mismatch condition being perceived as the least confident (see **Table S4**). In the 12 channel condition, there was no significant difference in the mean confidence rating between low and medium entropy sentences but participants were more confident in both these conditions compared to high and low entropy (mismatch) sentences. In the 1 channel condition, participants were more confident about the response after a low entropy sentence compared to a medium, high and low entropy mismatch sentence. After medium entropy sentences, participants were more confident than after high entropy sentences, however there was no difference in the mean confidence rating between medium and low entropy mismatch sentence. Finally, participants were significantly more confident after low entropy mismatch sentences in the 1 channel condition than after high entropy sentences.

Pearson correlation analysis between subjective clarity ratings and subclinical and cognitive trait markers are depicted in **Figure S2**.

| **Table S3: Results of the repeated measures ANOVA investigating the effect of condition and channel number on the mean confidence rating** | | | | | |
| --- | --- | --- | --- | --- | --- |
| **Effect** | **df** | **MSE** | **F** | **ges** | **p.value** |
| condition | 1.92, 90.14 | 193.98 | 245.08 *** | .360 | <.001 |
| channel_nr | 1.48, 69.75 | 444.73 | 598.96 *** | .709 | <.001 |
| condition:channel_nr | 5.66, 265.83 | 113.07 | 42.30 *** | .143 | <.001 |

| **Table S4: Results of Tukey post hoc tests investigating the interaction effect of entropy condition and channel number on the mean subjective confidence rating** | | | | | | |
| --- | --- | --- | --- | --- | --- | --- |
| **contrast** | **channel number** | **estimate** | **SE** | **df** | **t-ratio** | **p-value** |
| el - em | 12 | 0.73 | 0.57 | 47 | 1.28 | 0.58 |
| el - eh | 12 | 4.93 | 0.75 | 47 | 6.59 | <.001 |
| el - elmm | 12 | 21.01 | 1.98 | 47 | 10.59 | <.001 |
| em - eh | 12 | 4.21 | 0.79 | 47 | 5.31 | <.001 |
| em - elmm | 12 | 20.29 | 1.86 | 47 | 10.90 | <.001 |
| eh - elmm | 12 | 16.08 | 2.05 | 47 | 7.86 | <.001 |
| el - em | 6 | 2.43 | 0.73 | 47 | 3.34 | 0.01 |
| el - eh | 6 | 12.52 | 1.52 | 47 | 8.23 | <.001 |
| el - elmm | 6 | 35.88 | 2.23 | 47 | 16.08 | <.001 |
| em - eh | 6 | 10.09 | 1.29 | 47 | 7.83 | <.001 |
| em - elmm | 6 | 33.45 | 2.00 | 47 | 16.74 | <.001 |
| eh - elmm | 6 | 23.37 | 2.05 | 47 | 11.40 | <.001 |
| el - em | 3 | 8.92 | 1.51 | 47 | 5.92 | <.001 |
| el - eh | 3 | 29.28 | 1.70 | 47 | 17.27 | <.001 |
| el - elmm | 3 | 42.34 | 2.09 | 47 | 20.22 | <.001 |
| em - eh | 3 | 20.36 | 2.08 | 47 | 9.80 | <.001 |
| em - elmm | 3 | 33.42 | 2.41 | 47 | 13.89 | <.001 |
| eh - elmm | 3 | 13.06 | 1.72 | 47 | 7.60 | <.001 |
| el - em | 1 | 10.31 | 1.69 | 47 | 6.09 | <.001 |
| el - eh | 1 | 22.62 | 2.56 | 47 | 8.85 | <.001 |
| el - elmm | 1 | 13.74 | 2.61 | 47 | 5.27 | <.001 |
| em - eh | 1 | 12.32 | 1.99 | 47 | 6.20 | <.001 |
| em - elmm | 1 | 3.44 | 2.20 | 47 | 1.56 | 0.41 |
| eh - elmm | 1 | -8.88 | 2.43 | 47 | -3.65 | <.001 |

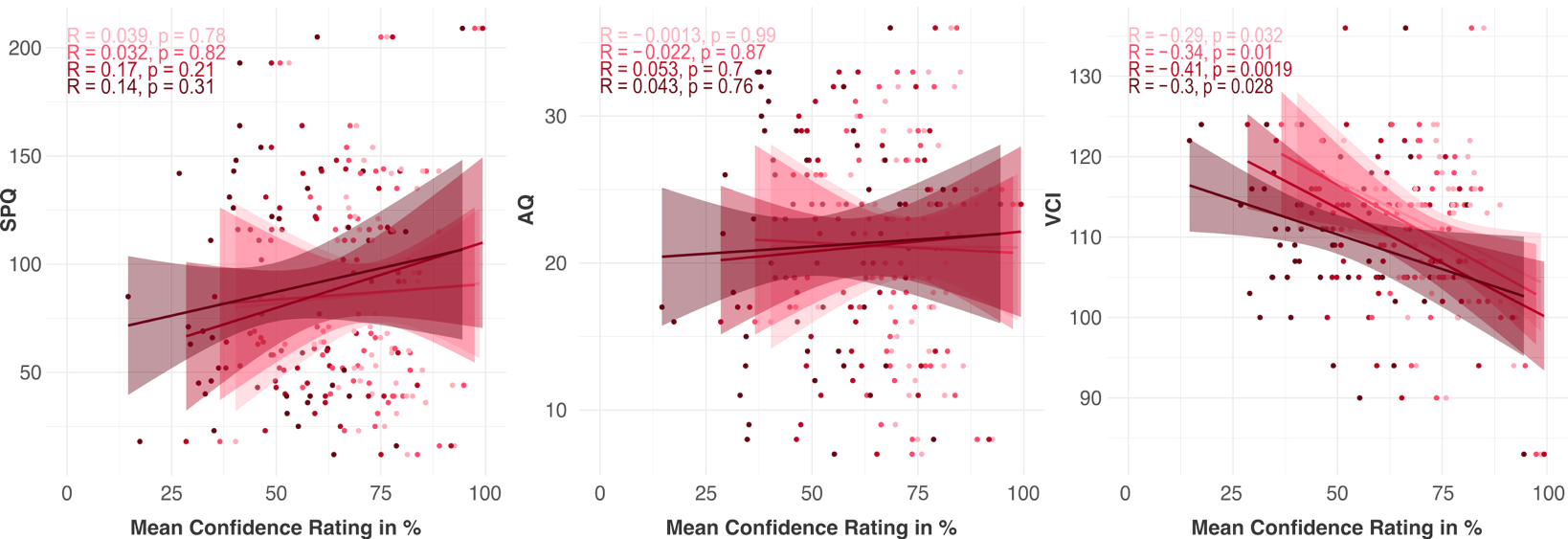

**Figure S2.** Scatter plots between subclinical and cognitive trait markers and subjective confidence ratings. P-Values are uncorrected.

R: Pearson Correlation Coefficient; SPQ: Schizotypal Personality Questionnaire(Raine, 1991; Wuthrich & Bates, 2005); AQ: Autism Spectrum Quotient(Baron-Cohen et al., 2001); VCI: Verbal Comprehension Index derived from the WAIS-IV(Wechsler, 2012)

**Statistical results showing the impact of sentence entropy behavioural response types**

To investigate the effect of entropy condition on the behavioural response types, repeated-measures ANOVAS were conducted separately for correct responses, no responses, and task-based hallucinations. For the proportion of correct responses, Mauchly’s test indicated that the assumption of sphericity had been violated (W = 0.65, p < .001, ε = .80). Therefore, degrees of freedom were corrected using Greenhouse-Geisser estimates of sphericity. There was a significant main effect of entropy condition (F(2.39, 129.24) = 553.00, p<0.001, general effect size = .852) on the proportion of correct responses. Tukey post hoc tests revealed a graded effect of the entropy condition on the proportion of correct responses with the highest proportion of correct responses in the low entropy condition, followed by the medium and high entropy condition, with the low entropy mismatch condition showing the lowest proportion (see **Table S5**). Pearson correlation analyses between the correct responses and subclinical and cognitive trait markers are depicted in **Figure S3**.

| **Table S5: Results of Tukey post hoc tests investigating the effect of entropy condition on the proportion of correct responses** | | | | | |
| --- | --- | --- | --- | --- | --- |
| contrast | estimate | SE | df | t-ratio | p-value |
| el - em | 8.70 | 0.76 | 54 | 11.51 | <.001 |
| el - eh | 26.95 | 0.95 | 54 | 28.29 | <.001 |
| el - elmm | 40.05 | 1.30 | 54 | 30.81 | <.001 |
| em - eh | 18.25 | 1.05 | 54 | 17.33 | <.001 |
| em - elmm | 31.34 | 1.22 | 54 | 25.64 | <.001 |
| eh - elmm | 13.09 | 1.13 | 54 | 11.63 | <.001 |

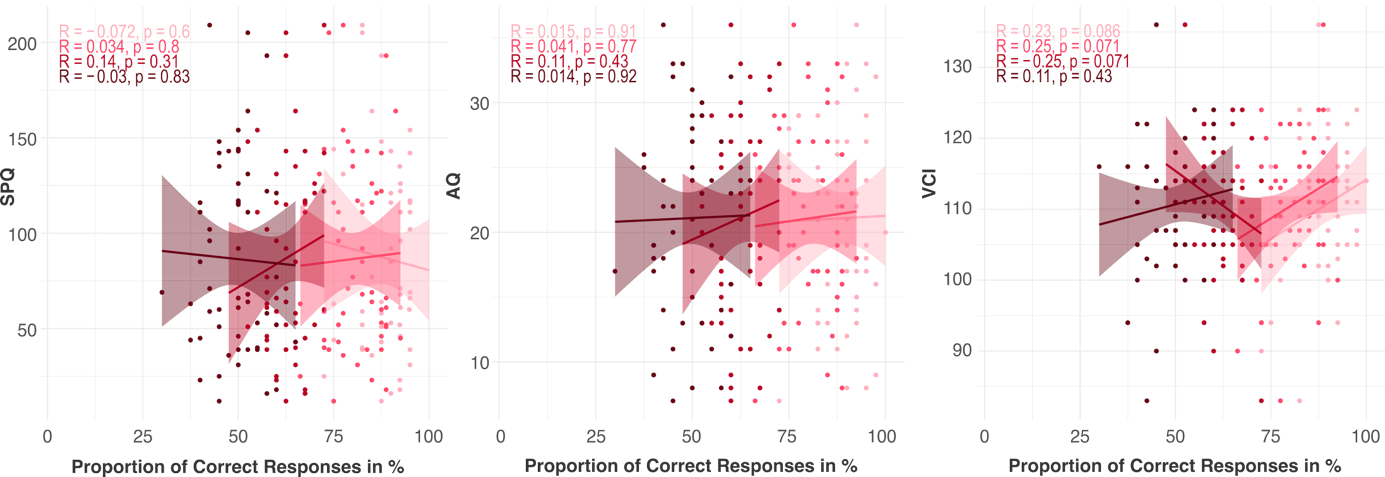

**Figure S3.** Scatter plots between subclinical and cognitive trait markers and correct responses. P-Values are uncorrected.

R: Pearson Correlation Coefficient; SPQ: Schizotypal Personality Questionnaire(Raine, 1991; Wuthrich & Bates, 2005); AQ: Autism Spectrum Quotient(Baron-Cohen et al., 2001); VCI: Verbal Comprehension Index derived from the WAIS-IV(Wechsler, 2012)

For the proportion of no responses, Mauchly’s test indicated that the assumption of sphericity had been violated (W = 0.44, p < .001, ε = .75). Therefore, degrees of freedom were corrected using Greenhouse-Geisser estimates of sphericity. There was a significant main effect of entropy condition (F(2.25, 121.29)=64.98, p<0.001, eta=0.199) on the proportion of no responses. Tukey post hoc tests revealed a graded effect of the entropy condition on the proportion of no responses with the highest proportion of no responses in the high entropy condition, followed by the low entropy mismatch and medium entropy condition, with the low entropy condition showing the lowest proportion (see **Table S6**). Pearson correlation analyses between the no responses and subclinical and cognitive trait markers are depicted in **Figure S4**.

| **Table S6: Results of Tukey post hoc tests investigating the effect of entropy condition on the proportion of no responses** | | | | | |
| --- | --- | --- | --- | --- | --- |
| contrast | estimate | SE | df | t.ratio | p.value |
| el - em | -3.75 | 0.63 | 54 | -5.92 | <.001 |
| el - eh | -13.55 | 1.24 | 54 | -10.95 | <.001 |
| el - elmm | -8.50 | 1.19 | 54 | -7.13 | <.001 |
| em - eh | -9.80 | 1.04 | 54 | -9.38 | <.001 |
| em - elmm | -4.75 | 0.87 | 54 | -5.49 | <.001 |
| eh - elmm | 5.05 | 1.08 | 54 | 4.65 | <.001 |

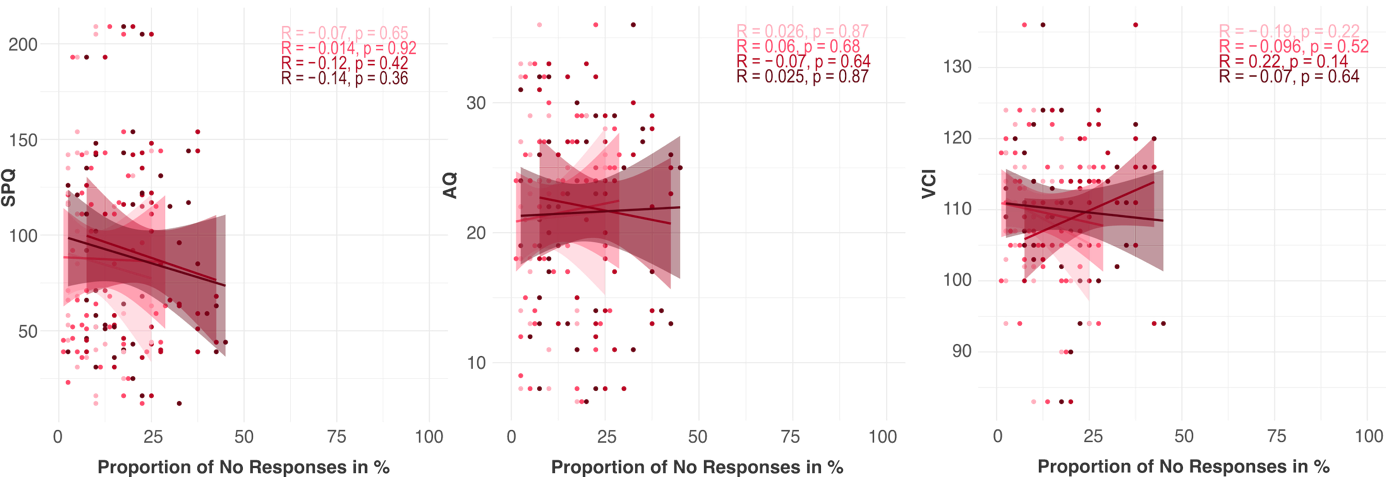

**Figure S4.** Scatter plots between subclinical and cognitive trait markers and no responses. P-Values are uncorrected.

R: Pearson Correlation Coefficient; SPQ: Schizotypal Personality Questionnaire(Raine, 1991; Wuthrich & Bates, 2005); AQ: Autism Spectrum Quotient(Baron-Cohen et al., 2001); VCI: Verbal Comprehension Index derived from the WAIS-IV(Wechsler, 2012)

For the proportion of task-based hallucinations, Mauchly’s test indicated that the assumption of sphericity had been violated (W = 0.30, p < .001, ε = .63). Therefore, degrees of freedom were corrected using Greenhouse-Geisser estimates of sphericity. There was a significant main effect of entropy condition (F(1.89, 102.05) = 199.22, p<0.001, general effect size = 0.617) on the proportion of no responses. Tukey post hoc tests revealed a graded effect of the entropy condition on the proportion of task-based hallucinations with the highest proportion of task-based hallucinations in the low entropy mismatch condition, followed by the high and medium entropy condition, with the low entropy condition showing the lowest proportion (see **Table S7**).

| **Table S7: Results of Tukey post hoc tests investigating the effect of entropy condition on the proportion of task-based hallucinations** | | | | | |
| --- | --- | --- | --- | --- | --- |
| contrast | estimate | SE | df | t-ratio | p-value |
| el - em | -4.95 | 0.67 | 54 | -7.35 | <.001 |
| el - eh | -13.41 | 1.42 | 54 | -9.47 | <.001 |
| el - elmm | -31.55 | 1.83 | 54 | -17.23 | <.001 |
| em - eh | -8.45 | 1.20 | 54 | -7.04 | <.001 |
| em - elmm | -26.59 | 1.57 | 54 | -16.92 | <.001 |
| eh - elmm | -18.14 | 1.36 | 54 | -13.31 | <.001 |

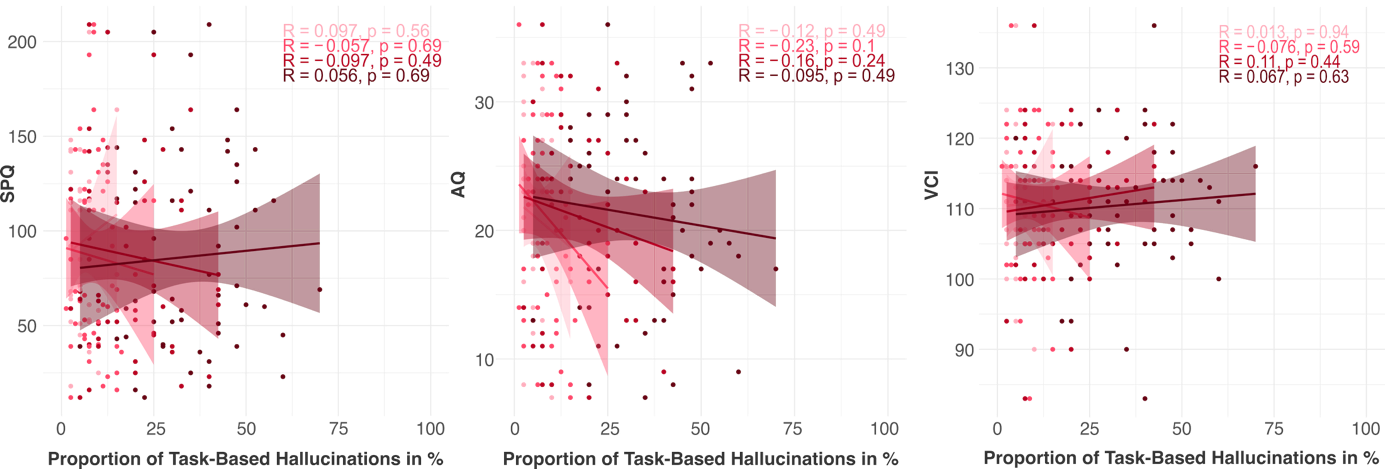

**Figure S5.** Scatter plots between subclinical and cognitive trait markers and task-based hallucinations. P-Values are uncorrected.

R: Pearson Correlation Coefficient; SPQ: Schizotypal Personality Questionnaire(Raine, 1991; Wuthrich & Bates, 2005); AQ: Autism Spectrum Quotient(Baron-Cohen et al., 2001); VCI: Verbal Comprehension Index derived from the WAIS-IV(Wechsler, 2012)

**Computational modelling – Quality checks**

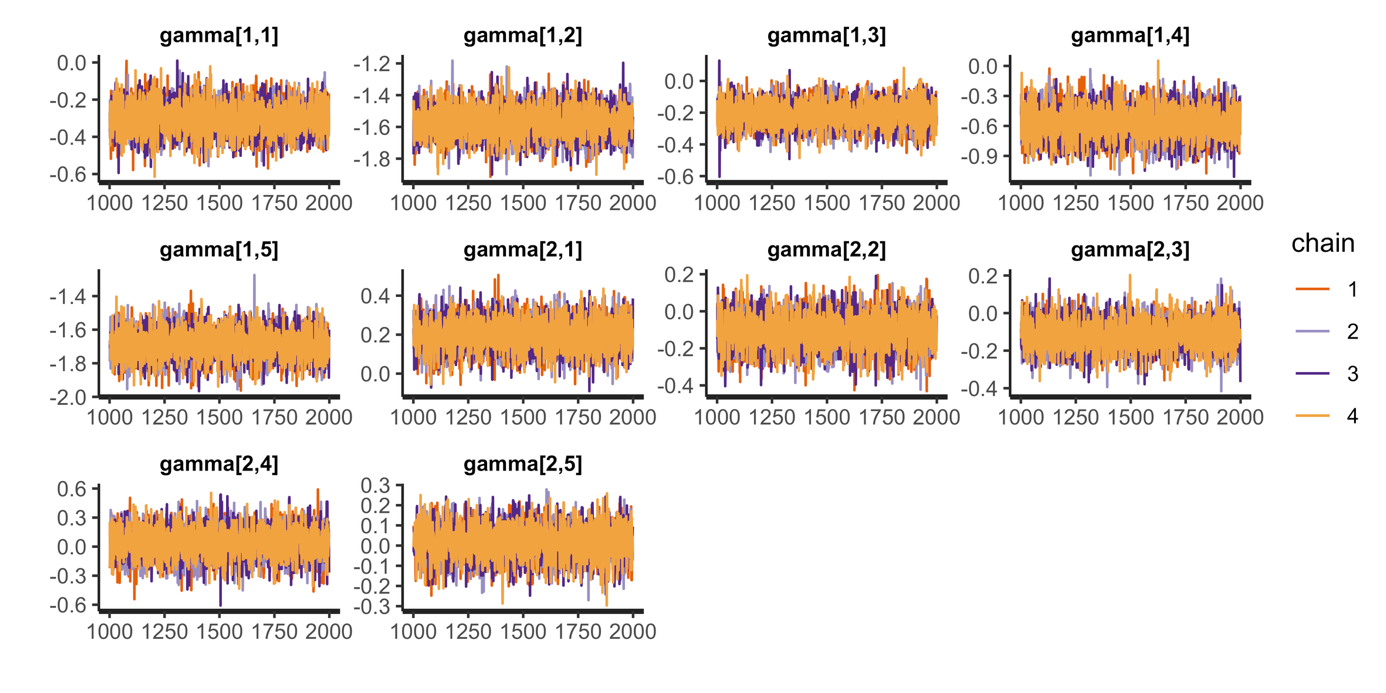

**Figure S6:** Trace plots for direct-effect model with individual effects for 55 participants for each population gamma showing successful convergence of four Markov Chains, each with 2000 iterations (warmup=1000; thin=1; post-warmup draws per chain=1000, total post-warmup draws=4000 for each gamma).

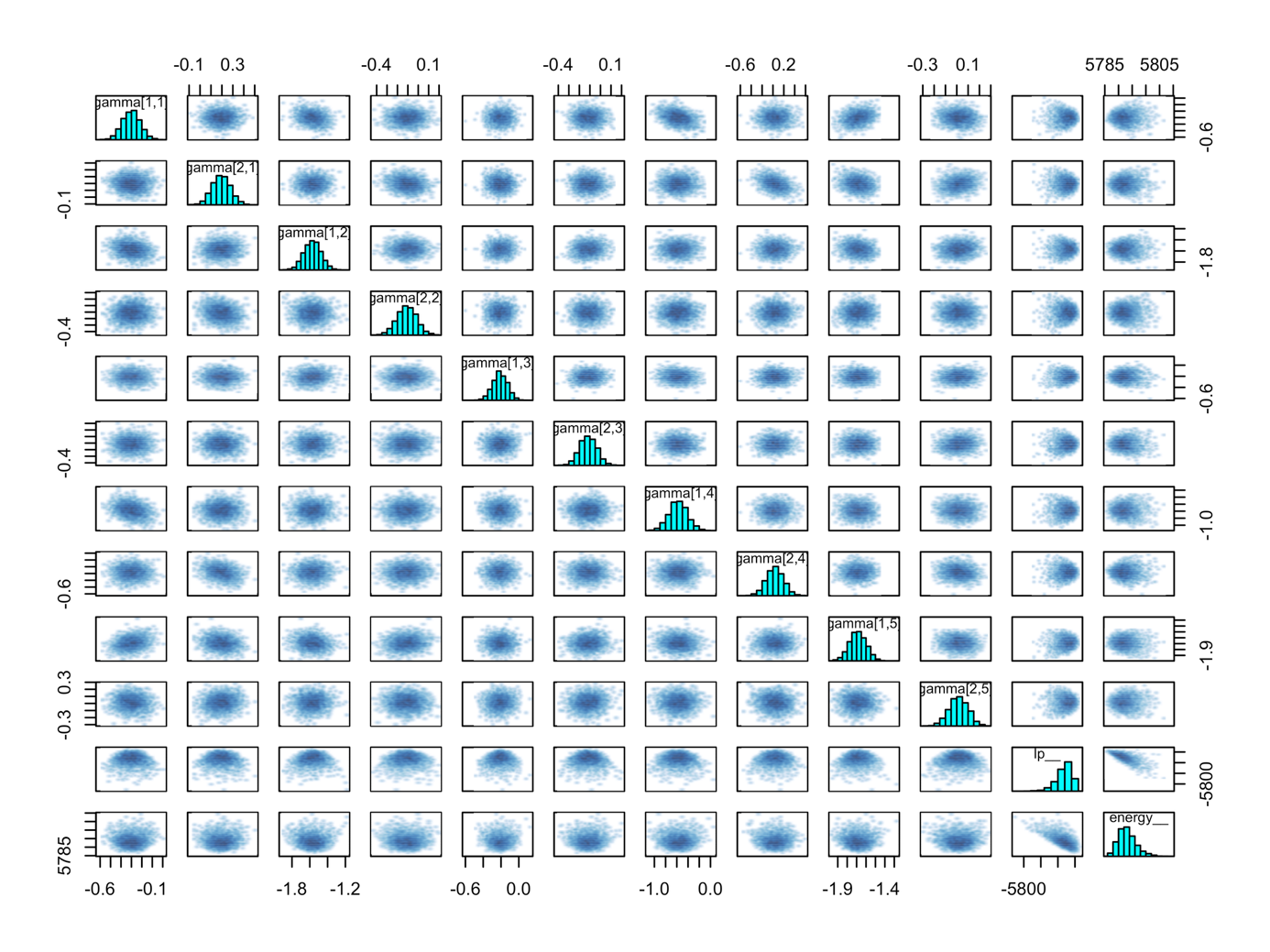

**Figure S7**: Pairwise scatterplots and marginal posterior distributions of population gammas. Diagonal histograms indicate normal distributions for all gammas. Scatterplot indicate little to no correlations between gammas.

**P600 Control Analysis**

| **Table S8: P600 LMM Results** | | | | | | | | | |
| --- | --- | --- | --- | --- | --- | --- | --- | --- | --- |
|  | **SPQ** | | | **AQ** | | | **VCI** | | |
| *Predictors* | *Estimates* | *CI (95%)* | *P-Value* | *Estimates* | *CI (95%)* | *P-Value* | *Estimates* | *CI (95%)* | *P-Value* |
| Intercept | 3.65 | 2.52 –  4.78 | **<0.001** | 3.65 | 2.52 –  4.78 | **<0.001** | 3.65 | 2.52 –  4.78 | **<0.001** |
| Medium Entropy | -0.05 | -0.22 –  0.12 | 0.570 | -0.05 | -0.22 –  0.12 | 0.570 | -0.05 | -0.22 –  0.12 | 0.568 |
| High Entropy | -1.00 | -1.17 –  -0.82 | **<0.001** | -1.00 | -1.17 –  -0.82 | **<0.001** | -1.00 | -1.17 –  -0.82 | **<0.001** |
| Low Entropy (Mismatch) | -1.98 | -2.16 –  -1.81 | **<0.001** | -1.98 | -2.16 –  -1.81 | **<0.001** | -1.98 | -2.16 –  -1.81 | **<0.001** |
| Trait Marker | 0.02 | -0.43 –  0.46 | 0.932 | 0.05 | -0.39 –  0.50 | 0.818 | 0.25 | -0.19 –  0.70 | 0.263 |
| Trait Marker: Medium Entropy | -0.23 | -0.40 –  -0.06 | **0.009** | 0.04 | -0.14 –  0.21 | 0.674 | 0.15 | -0.02 –  0.32 | 0.081 |
| Trait Marker: High Entropy | -0.11 | -0.28 –  0.06 | 0.221 | 0.08 | -0.10 –  0.25 | 0.383 | 0.02 | -0.15 –  0.19 | 0.813 |
| Trait Marker: Low Entropy (Mismatch) | -0.34 | -0.52 –  -0.17 | **<0.001** | -0.05 | -0.22 –  0.12 | 0.554 | -0.37 | -0.54 – -0.20 | **<0.001** |
| **Random Effects** | | | | | | | | | |
| σ^2^ | 3.18 | | | 3.19 | | | 3.16 | | |
| τ_00_ | 2.61 _participant_ | | | 2.63 _participant_ | | | 2.59 _participant_ | | |
|  | 4.20 _electrodeposition_ | | | 4.20 _electrodeposition_ | | | 4.19 _electrodeposition_ | | |
| ICC | 0.68 | | | 0.68 | | | 0.68 | | |
| N | 15 _electrodeposition_ | | | 15 _electrodeposition_ | | | 15 _electrodeposition_ | | |
|  | 55 _participant_ | | | 55 _participant_ | | | 55 _participant_ | | |
| Observations | 3300 | | | 3300 | | | 3300 | | |
| Marginal R^2^ / Conditional R^2^ | 0.065 / 0.702 | | | 0.062 / 0.701 | | | 0.069 / 0.704 | | |
